# Medical Cannabis Certifications for Severe Chronic and Intractable Pain: Discerning Geographic Patterns Across Pennsylvania, USA

**DOI:** 10.1101/2025.06.25.25329221

**Authors:** Sanya K. Mehta, Lorraine D. Tusing, Ahmad Higazy, Brian J. Piper

**Affiliations:** School of Medicine, University of Louisville, Louisville, KY; Department of Medical Education, Geisinger College of Health Sciences, Scranton, PA; Center for Pharmacy Innovation & Outcomes, Geisinger College of Health Sciences, Danville, PA

**Keywords:** medical cannabis, severe chronic and intractable pain

## Abstract

**Introduction:** Chronic pain is the most common qualifying condition found in states with medical cannabis (MC). We conducted a study assessing the geographic distribution of MC certifications for severe chronic or intractable pain in Pennsylvania (PA) between 2018 to 2024, identifying relationships between median household income and ethnic background with the percentage of adults with a MC certification for pain.

**Methods:** Using data from the PA Department of Health (PDOH) from 2018 to 2024 (N = 44,645 to 165,740 certifications for pain / year), we mapped Zip codes associated with MC certifications for pain to counties and Zip code tabulation areas (ZCTAs). The difference between the highest and lowest counties was determined. A linear regression evaluated correlations between community variables and the percentage of adults in geographical areas with a MC certification for pain in 2024.

**Results:** There was an almost a four-fold difference in the percent of adults with a MC certification for pain in the highest (Perry = 2.3%) versus lowest (Tioga = 0.6%) counties in 2024. Bradford and Tioga County had a significantly (p < 0.05) lower percentage certified relative to the county-wide average. There was a significantly higher proportion of certifications for pain in counties with larger population densities of adults (1.76 +/− 0.12%) than counties with smaller population densities (1.38% +/− 0.14%) of adults (t(65) = 4.66, p < 0.001, *d* = 1.14). At the county level, higher median household income was associated with a greater percentage of adults with MC (r(65) = +0.34, p < 0.01). At the ZCTA level, the proportion of non-White individuals, including Hispanics, showed a modest, but significant, inverse association with MC certification (r(1,722) = –0.07, p < 0.01).

**Conclusions:** This study identified four-fold county level disparities in MC certifications for pain. The association between median household income and MC pain certifications may indicate differences in accessibility of MC based on financial status. Further research may be warranted pending any changes to the legal status or demand for MC.

## Introduction

Access to medical cannabis (MC) has been increasing across the United States (US), with many states expanding their list of qualifying conditions [1]. Pennsylvania (PA) passed the Medical Marijuana Act (Act 16) in April 2016 to provide MC for patients with specified conditions [2]. Chronic pain was the most common qualifying condition reported by patients nationwide using MC [3]. Similarly, almost all (97%) of states with MC listed chronic pain as a qualifying condition [4]. The National Academy of Sciences rated the quality of evidence of cannabis for chronic pain in adults in 2017 as “substantial” [4]. Six randomized controlled trials support the use of low-dose MC in conjunction with traditional analgesics for refractory neuropathic pain [5]. However, there is limited knowledge of the long-term effects of MC usage for pain relief and the use of MC for other causes of chronic non-cancer pain, such as rheumatologic conditions [5]. An evidence map of systematic reviews completed though 2023 determined that the weight of the evidence of MC was generally “positive” for chronic pain, oncological pain, and pain relief but more divided between “potentially positive” and “positive” for neuropathic pain and “potentially negative” for post-operative pain [6]. As the utilization of MC expands across PA for chronic pain relief, it is crucial to study the geographic distribution and social determinants of MC utilization for pain to ensure that MC access is equitable and to inform MC policy and legislation.

The ability to access MC across the US is influenced by income, race/ethnic background, urban/rural residence, and stigma [7, 8]. The cost of MC may serve as a barrier to utilization, as health insurance does not currently cover the vast majority of cannabis products. According to the Pennsylvania Department of Health (PDOH), patients in the PA Medical Marijuana Program spend about $275 a month ($3,300/year) on MC [9]. In addition, a patient’s race or ethnic background may influence their likelihood to use MC for pain. Overall, MC use is less likely among non-White and foreign-born individuals [8]. Factors that may contribute to the underrepresentation of foreign-born individuals’ use of MC include exclusionary immigration policies that hinder participation in government programs and greater legal consequences of involvement with cannabis for minority groups [8]. Self-reported chronic pain was nearly sixty-percent greater among adults in suburban and rural communities compared to those in urban communities [10]. These urban-rural differences in chronic pain may be due to higher median age or rates of chronic disease in rural communities [11, 12].

To our knowledge, no research as of yet has examined the underlying geographic distribution of MC certifications for pain in PA and reported the associations with demographic factors. Our investigation leveraged MC certification data obtained from the PDOH to investigate these relationships.

## Methods

### Procedures

A retrospective study of MC certifications for severe chronic or intractable pain, including pain of neuropathic origin, (henceforth “pain”) in PA counties was conducted using data from the PDOH obtained from 2018-2024, geographic spatial files from the Census Bureau, and community sociodemographic data from the American Community Survey (ACS). One month of certification data (9/2020) was not reported by the PDOH (shown in Supplementary Fig. 1).

### MC Certifications at the Zip Code Level

For each year between 2018 and 2024, the PDOH provided certification data at the zip code level. Data included 5-digit zip codes, certification status (included: active, inactive, pending, expired, cancelled), creation date of certification, treatment period (by number of months up to 12), and up to ten qualifying serious medical conditions approved by the PDOH.

### Mapping Zip Codes to Counties and ZCTAs

Since zip code level analysis is often distorted due to the adverse statistical properties inherent with zip codes [13], we aggregated zip code data and summarized it to the county and ZCTA levels. Zip code to county crosswalk files obtained from the US Department of Housing and Urban Development (HUD) and county-level Gazetteer files were used to summarize MC certifications at the county level. Out of a total of 905,672 MC certifications from 2018 to 2024 for pain, 268,342 certifications were in zip codes that mapped to multiple counties. We assigned these to the county with the highest ratio of residences in that particular zip code to the total number residential addresses in the zip code (“res_ratio”) as reported in HUD crosswalks. We used this approach for our main analysis since some MC certifications for pain (≈ 0.5%) were in zip codes that mapped to counties with residential ratios of 0 [13]. The validity of this approach has been assessed in the literature [13]. In addition, we conducted a sensitivity analysis to ensure that we were not systematically excluding certain counties from our analysis and assess the validity of our results. In our sensitivity analysis, we used proportional allocation to map certifications: certification frequencies by zip code were weighted by the corresponding residential ratio (“res_ratio”) from the HUD crosswalk (Supplementary Fig. 2; Supplementary Fig. 3).

Since minors account for a small fraction (≈ 0.2%) of active MC certifications as of November 2024 [14], we divided the number of certifications in each year between 2018 and 2023 by the size of the adult population in that county using yearly data from the ACS. This practice has previously been reported in the literature to assess the geographical distribution of MC certifications in PA [15]. Since 2024 ACS data was not publicly available at the time of this study, we divided the number of certifications in 2024 by the size of the adult population in that geographical unit in 2023.

Zip code to ZCTA crosswalk files from a GitHub repository from UDS Mapper (2018-2022) and from HRSA (2023), using March 2023 boundaries for zip codes and the Census Tiger 2022 geographic boundaries for ZCTAs, were used to summarize MC certifications at the ZCTA level [16, 17]. Since a zip code to ZCTA crosswalk was not available for 2024, the 2023 crosswalk mappings were carried forward. ZCTAs that had estimated adult population sizes lower than the number of certifications mapped to that ZCTA were removed.

### Analysis

The goals of these analyses were to describe and map the geographical distribution of MC certifications in PA; assess the correlations between two community variables: median household income and the percent of adult individuals identifying as non-white, including those of Hispanic ethnicity; map regions of low-use and high-use of MC for pain, along with spatial outliers; and assess the spatial autocorrelation of the mapped distribution of the percentage of adults certified for MC for pain.

We conducted Moran analyses of the percent of adults with a MC certification for pain from 2018 to 2024 using Queen-based spatial neighbors without using a model or offset at the county and ZCTA levels (Supplementary Table 1). In addition, we mapped local Moran’s I patterns based on the proportion of adults with an MC certification for pain to identify geographical clusters and spatial outliers. Spatial units were characterized as hotspots, coldspots, high-low spatial outliers, or low-high spatial outliers if local p-values were less than or equal to 0.05 (Supplementary Fig. 4; Supplementary Fig. 5). We conducted an additional analysis at the county level for which we applied a statistical correction using the Benjamini-Hochberg procedure and mapped clusters and outliers if adjusted local p-values were less than or equal to 0.05 (Supplementary Fig. 6).

We utilized linear regression to analyze the correlations between 1) median household income and 2) the percentage of adult (≥ 18 years) individuals identified as non-White, including those of Hispanic ethnicity, and percentages of adults with cannabis certifications for pain in each geographical unit in 2024 at the county and ZCTA levels. Again, since 2024 ACS data was not publicly available at the time of this study, we divided the number of certifications in 2024 by the size of the adult population in that geographical unit in 2023 and analyzed the correlation of this statistic with 2023 ACS demographic variables.

The PA counties with a corresponding percentage of adults with cannabis certifications for pain greater than ± 1.96 standard deviations (SDs) from the county-wide mean were considered significant although other counties that met a more liberal threshold (1.5 SD) were also noted. Counties were split by median adult population (68,169) into a high population group (N = 34) or low population group (N = 33). In a follow-up analysis, counties were split by median population density of adults using the same procedure (Supplementary Fig. 7). A Student’s two-sample t-test assuming equal variances compared the percent of a county population certified for pain by adult population.

The degrees of freedom of t-tests and correlation coefficients were recorded for all analyses. Variability was represented using the standard deviation from the mean. Significant group differences were expressed in terms of Cohen’s *d* where 0.2, 0.5, 0.8, and 1.3 are interpreted as a small, medium, large, and very large (respectively) effect size [18]. All analyses were conducted in R version 4.4.0. Geographical boundaries were outlined using the tigris package, spatial analyses were conducted using the sdep package, and mapping was performed using ggplot2. The multivariable negative binomial regression models were conducted using the glm.nb function in the R package MASS.

## Results

Out of a total of 905,672 MC certifications, we were able to map 905,590 (99.99%) to a county using a one-to-one mapping technique mapping each zip code to the corresponding county with the highest residential representation (shown in Fig. 1). Further, we mapped 905,599 (99.99%) certifications to one of the 1,831 ZCTAs (Supplementary Fig. 8).

**Fig. 1.**
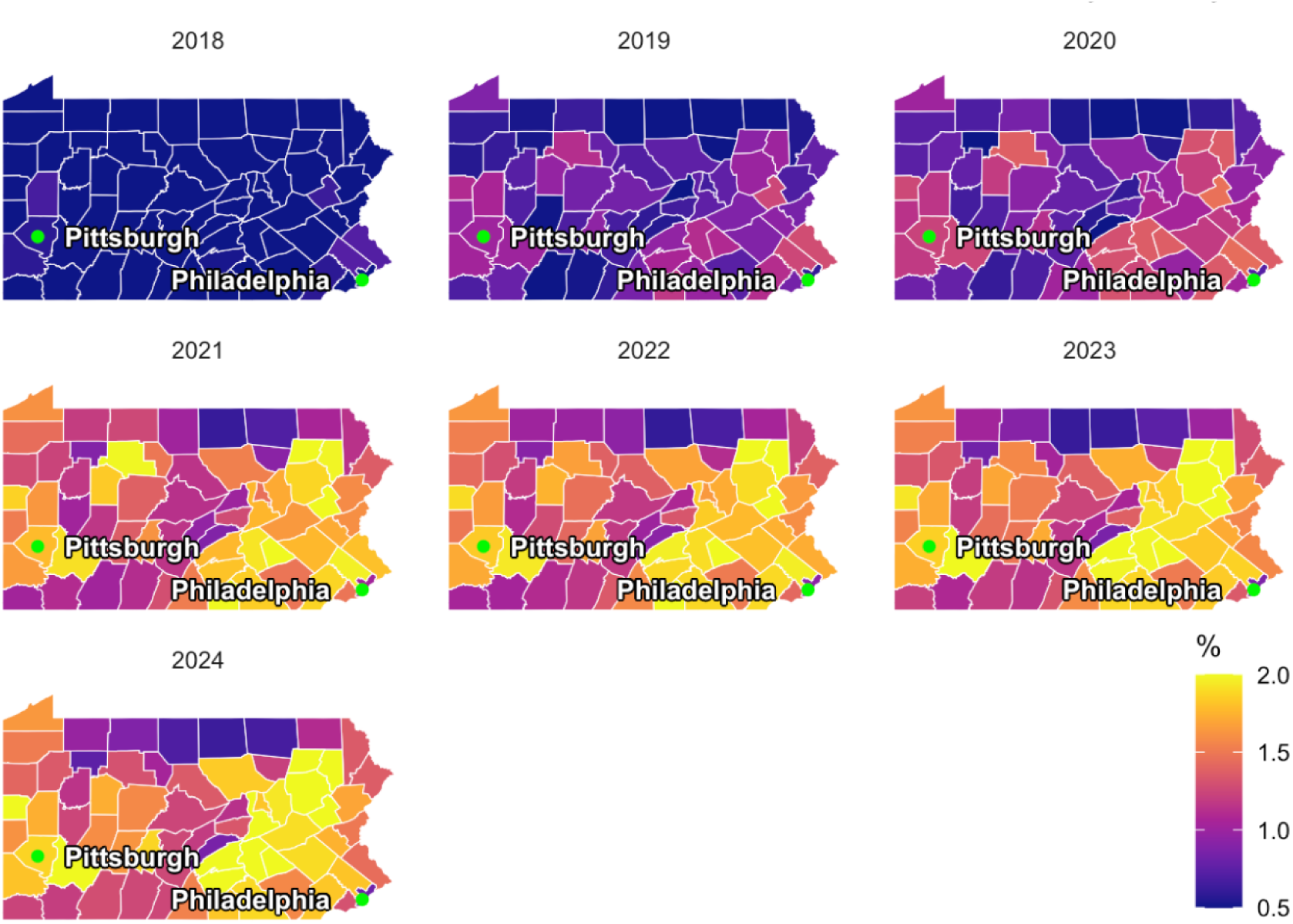
Heat map of the percent of adults in Pennsylvania with a medical cannabis certification for pain from 2018–2024 by county.

Overall, counties with larger adult populations had greater MC certification rates (1.74 +/− 0.11%) than smaller (1.31 +/− 0.15%) counties (t(65) = 4.72, p < 0.001, *d* = 1.15). In addition, counties with larger population densities of adults (1.76 +/− 0.12%) had greater MC certification rates than counties with smaller (1.38 +/− 0.14%) population densities (t(65) = 4.66, p < 0.001, *d* = 1.14, Supplementary Fig. 7). Bradford and Tioga Counties had a significantly lower percent of adults with a MC certification for pain (p < 0.05) relative to the average across all counties (1.53%) in 2024 (Fig. 2). Consistent results were observed in our sensitivity analysis (Supplementary Fig. 2; Supplementary Fig. 3).

**Fig. 2.**
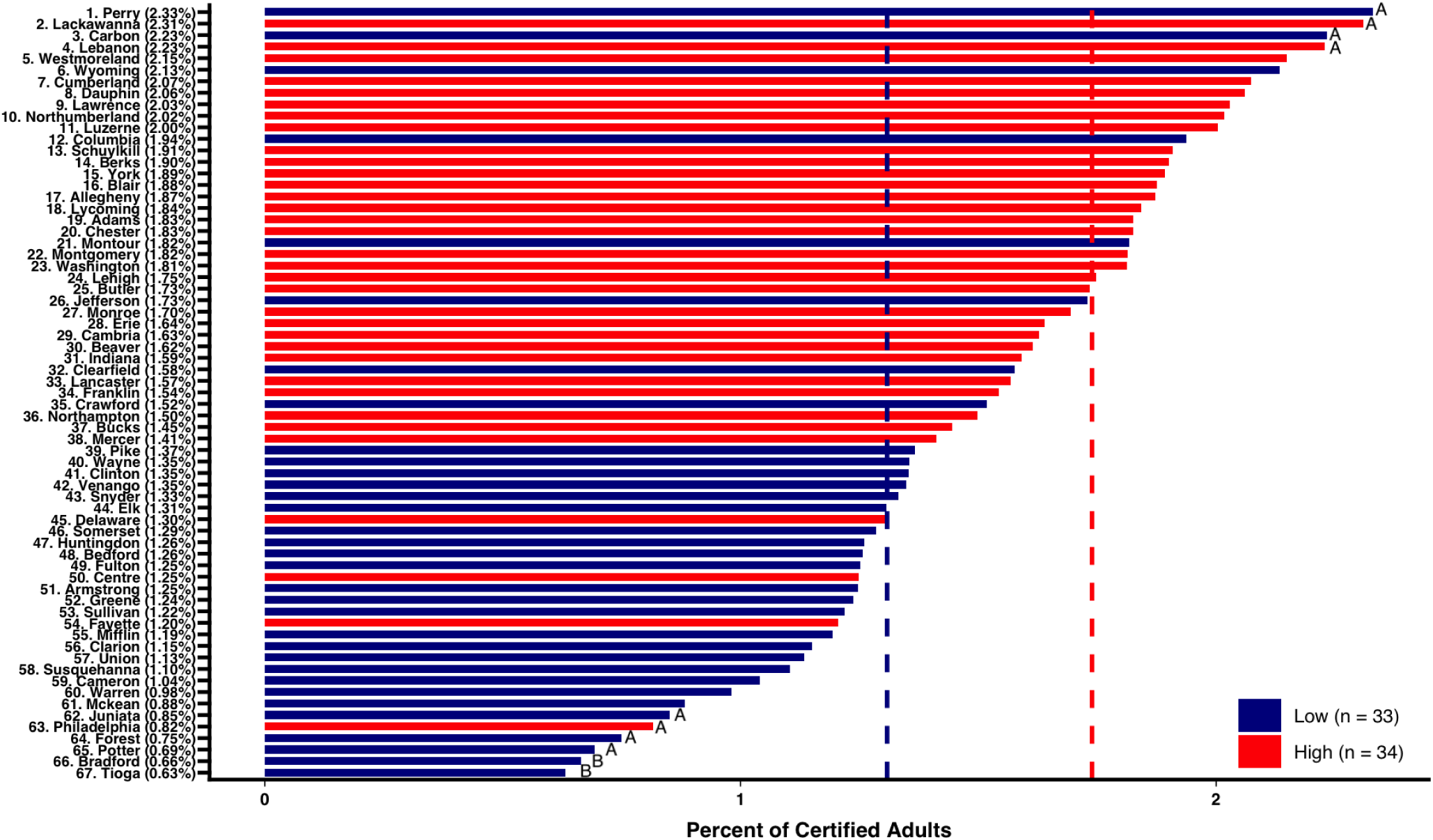
Waterfall plot of the percent of adults certified for medical cannabis for pain in Pennsylvania by county and by population size. Counties with a Z score > ±1.50^A^ or > ±1.96^B^.

There was a significant positive association between median household income and the proportion of adult residents with a MC certification for pain in 2024 at the county level (r(65) = +0.335, p = 0.00552, Fig. 3). After removing counties with an associated median household income higher than $100,000, the significant correlation was retained (r(62) = +0.400, p = 0.00107). In addition, there was a small but significant negative correlation between the percent of non-White individuals, including those of Hispanic ethnicity, and the proportion of adult residents with a MC certification for pain at the ZCTA level in 2024 (r(1,722) = –0.0749, p = 0.001885, Supplementary Fig. 9). There was no significant relationship in 2024 between the percent of non-White individuals, including Hispanics, and the proportion of adult residents with a MC certification at the county level (r(65) = 0.142, p = 0.2505, Supplementary Fig. 10) and no significant relationship between median household income and the proportion of adult residents with a MC certification at the ZCTA level (r(1,606) = 0.0278, p = 0.264, Supplementary Fig. 11). For reference, we have also mapped the county-level distribution of adult population and median household income (Supplementary Fig. 12; Supplementary Fig. 13).

**Fig. 3.**
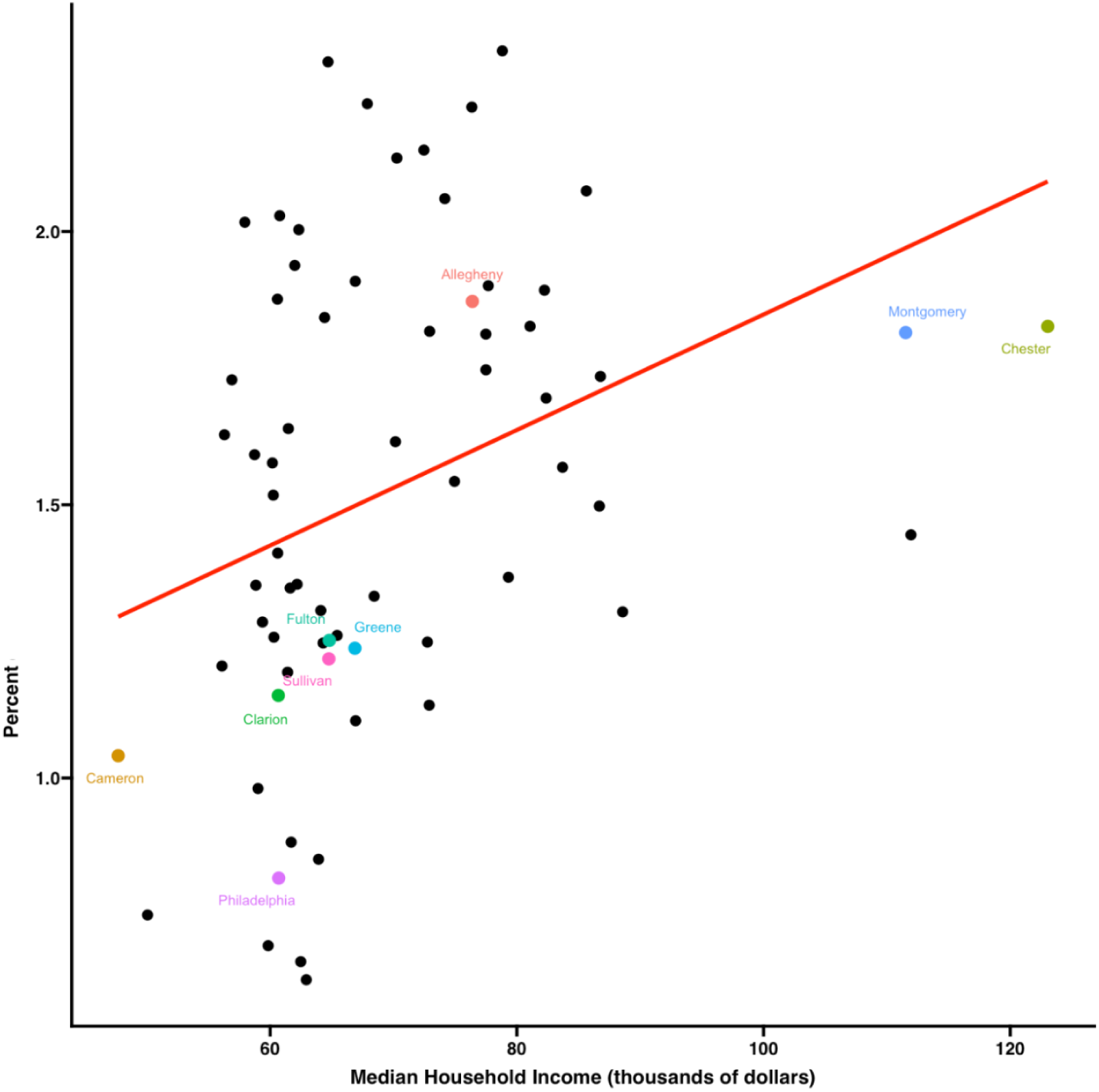
Scatterplot of the percent of adults with a MC certification for pain versus median household income in 2024 at the county level. (r(65) = +0.335, p = 0.00552).

## Discussion

As MC becomes increasingly available, it is essential to study the geographical distribution and identify factors that influence access to MC for the treatment of pain. We conducted the first analysis of geographical patterns and barriers to access of utilization of MC for pain. In line with the findings of the State Epidemiological Outcomes Workgroup in 2022 reporting the patient density of total MC certifications per county for all indications as of August 2021, we found that Bradford County and Tioga County also had a significantly lower proportion of adults with a MC certification for pain as compared to the county-wide average in 2024 [19]. The four-fold disparities between the highest and lowest counties may be concerning because chronic pain in adults is one of the few qualifying medical conditions in PA where the 2017 NAS report rated the quality of the evidence base that cannabis is effective as “substantial” [4].

Although there has been little research on the associations between sociodemographic variables and the frequency of MC certifications for pain, previous reports analyzing geographic access to MC services across states have found mixed results regarding the influence of median household income on MC accessibility [20–23]. While prior studies of access to MC dispensaries in California and Colorado found no significant association between income and geographic access to MC dispensaries, a report in Oklahoma found that higher proportions of residents who were uninsured and living in poverty were associated with less availability of MC services [20–22]. In contrast, an investigation from New York found that MC services were least available in neighborhoods with highly educated residents [23]. Our study found a significant correlation between the percent of non-White individuals including those of Hispanic ethnicity and the proportion of adults with a MC certification for pain at the ZCTA level.

From an epidemiological perspective where those residing in rural areas are older, more likely to work in manufacturing or agricultural jobs, and more likely to report chronic pain [24], one might anticipate that MC for pain would be higher in rural vs urban areas. However, marijuana is currently a Schedule I substance so MC is not covered by insurance. Chronic pain patients in New England spent a considerable amount (median = $2,320, mean = $3,064) of their disposable income on MC [25]. Dispensaries tend to be localized in more urban areas [15]. In our analysis, we found a significant positive correlation between median household income and the proportion of adults with MC certifications for pain in 2024 at the county level. This suggests that patients’ propensity to utilize MC for pain relief may be influenced by their median household income in addition to their geographical location. The cost of a high-quality ounce of MC in PA ($384) was well above the national average ($326) and ranked seventh in the US [26]. The annual cost ($50) for registering for a medical marijuana identification card can be waived for those enrolled in Medicaid, Children’s Health Insurance Program, Supplemental Nutrition Assistance Program, and the Supplemental Nutrition Program for Women, Infants, and Children [27]. As such, our findings suggest that it could be beneficial for primary care providers to consider the ability of patients to afford medical cannabis and geographic barriers to utilization when incorporating MC into the treatment plan of patients with severe chronic or intractable pain.

There is a long and unfortunate racialized history in this field with some perceiving the term “marijuana” as racist [28]. Doctors from a large health system in western PA were almost three-fold more likely to test a Black than a White pregnant woman for marijuana and other drugs despite the urine toxicology being less likely to be positive for Black than White patients [29]. A Black person was three times more likely to be arrested for marijuana possession than a White person in PA [30]. The disparity was even larger in 2018 in Clarion (25.6 fold) and Parry (28.4 fold) counties [30]. Nationally, past month marijuana use among those greater than or equal to 12 years of age were similar among Black (18.1%) and White (16.3%) people [31]. Although pain is a biopsychosocial condition, reporting pain severity is subjective. We can not discount the possibility that some patients obtain a MC certification for pain or other medical conditions [15] in an attempt to minimize their legal risk. Although we found that ZCTAs with greater non-White populations had significantly fewer pain certifications, the percentage of variance accounted for (0.6%) was so modest as to be practically meaningless [18].

There are some caveats to this report. One limitation of this analysis is that the de-identified data was submitted to us from a third-party source (Supplementary Fig. 1). There are multiple versions of MC certification datasets as the PDOH has updated the data several times. We anticipate that it is likely that the PDOH will make amendments to the datasets that were provided to us [14]. Another limitation is the potential for confounding, as counties with higher median household incomes and higher proportions of non-White individuals may differ in other relevant ways—such as adult population size, provider density, and healthcare access —that could influence certification rates. An additional limitation includes the potential of zip codes to map to multiple geographic units, leading to inaccuracies in mapping. In addition, ACS estimates at the local level are often less reliable with larger margins of error which may significantly influence the results of our ZCTA-level analysis [32]. This investigation was completed in one state (PA) and assessed medical but not recreational cannabis. These findings may not generalize to other states with different MC policies [28]. A prior report from a resource limited population in the Bronx determined that over half (55%) of newly certified patients, predominantly for pain, did not purchase MC [33]. We do not know how many of the certified patients in PA were regularly using MC. Additional research is necessary to examine whether the population size or income geographical differences also apply to illicit cannabis or to delta-8 tetrahydrocannabinol use [34].

In conclusion, we have mapped the geographic distribution of the percent of adults with a MC certification for pain in PA from 2018 to 2024 at the county and ZCTA levels. We found approximately a four-fold difference in the percent of adults with a MC certification for pain between the county with the highest and lowest certification rate in 2024. In addition, we report significant correlations between 1) income and 2) ethnicity and the percent of adults with a MC certification for pain in 2024.

## Acknowledgements

We would like to thank Annemarie Hirsch, PhD, for her valuable suggestions regarding statistical tests to run to strengthen the robustness of our analysis. We would also like to thank Ed Mahon for his efforts to make this data publicly available.

GPT-4 and GPT-4o [35] were used to assist in generating and debugging code sections utilized in the data-analysis. S.K.M. verified the accuracy of all generated code by checking the function of the code using R manuals (accessible at https://cran.r-project.org/), comparing the code outputs to manual counts of certification frequencies, and running test cases. S.K.M. also went through all generated summary tables and ensured their accuracy. Further, the authors assessed the robustness of the data to various mapping techniques, some of which are detailed in the supplementary material.

## Author Contributions

S.K.M. and B.J.P. were involved in the conception of the work and the design of the study; L.D.T. and B.J.P. were involved in the acquisition of the data; S.K.M. and A.H. were involved in the analysis of the data; S.K.M., A.H., and B.J.P. were involved in the interpretation of the data; S.K.M. drafted the manuscript; and all authors were involved in substantially revising the manuscript.

## Statement of Ethics

The Geisinger Institutional Review Board (GIRB) indicated that the study proposal did not meet the definition of *Research* as defined in 45 CFR 46.102(d): *a systematic investigation, including research development, testing, and evaluation, designed to develop or contribute to generalizable knowledge*. Therefore, this proposal was deemed not subject to human subjects’ research regulations and was exempt from oversight by GIRB and also exempt from requiring written informed consent.

## Data Availability Statement

The datasets generated and/or analyzed during the current study are not publicly available as the data provided regarding MC certifications were provided by a third-party source that requires approval of data use by the IRB designated to the Academic Clinical Research Center (Geisinger).

## Supplementary Material

**Supplementary Fig. 1.**
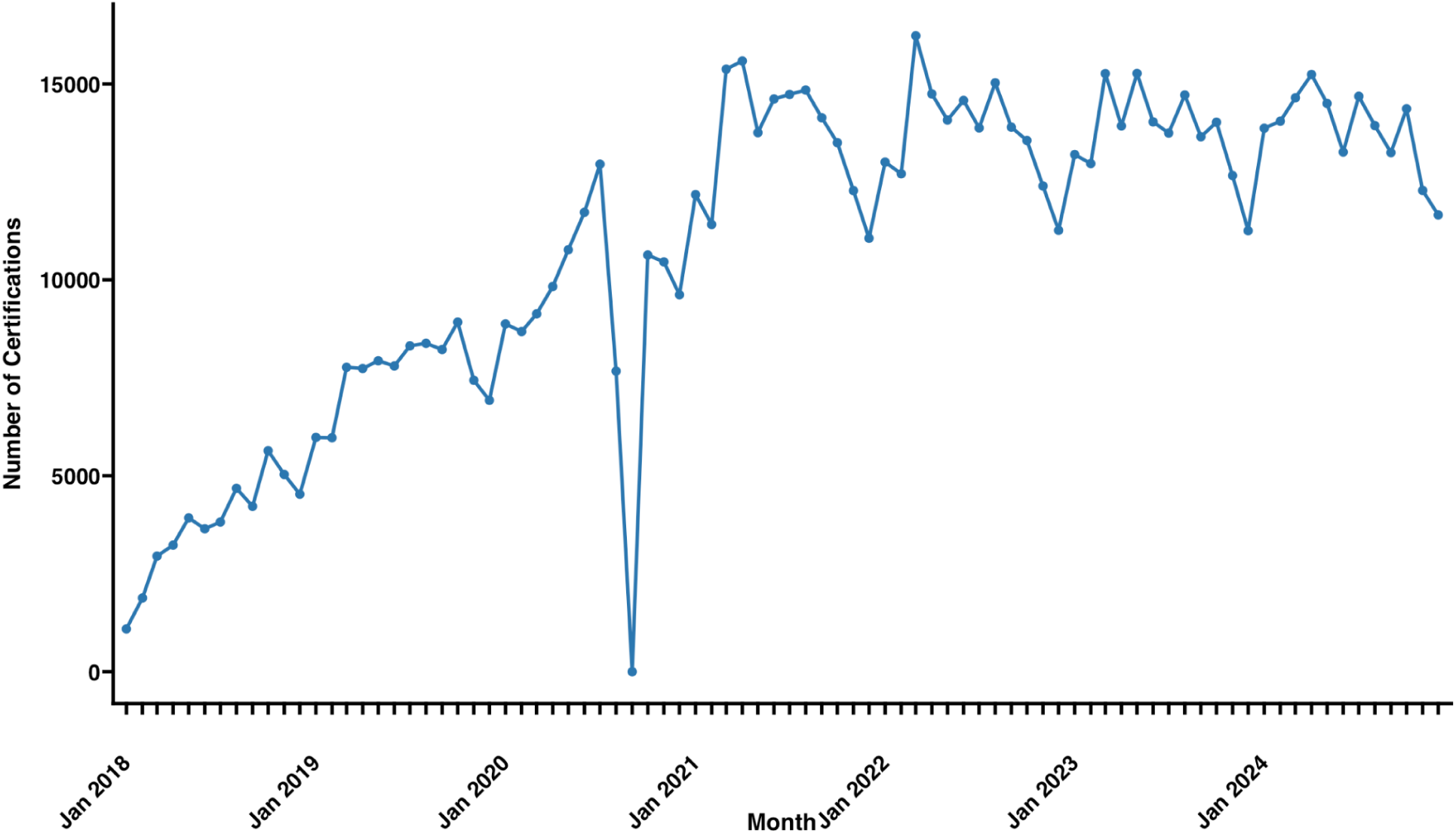
Monthly counts of MC certifications for pain. No MC certifications for pain were reported in September 2020.

**Supplementary Fig. 2.**
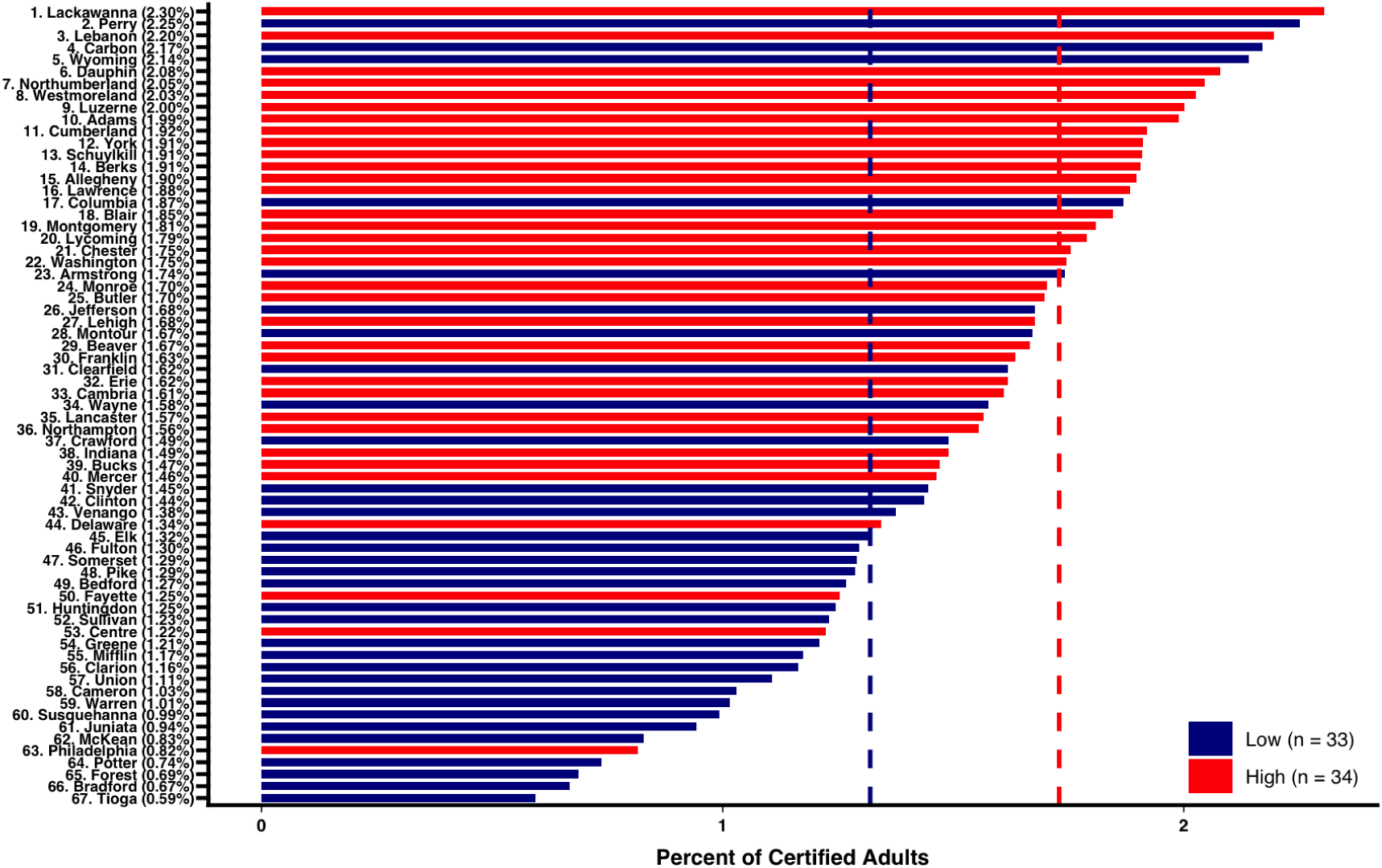
Waterfall plot of the percent of adults certified for medical cannabis for pain in Pennsylvania by county and by population size using proportional allocation of residential ratios. Using proportional allocation, there was a significantly higher proportion of MC certifications for pain in more populated (1.73 +/− 0.10%) than less populated (1.32% +/− 0.15%) counties (t(65) = 4.60, p < 0.001, *d* = 1.22).

**Supplementary Fig. 3.**
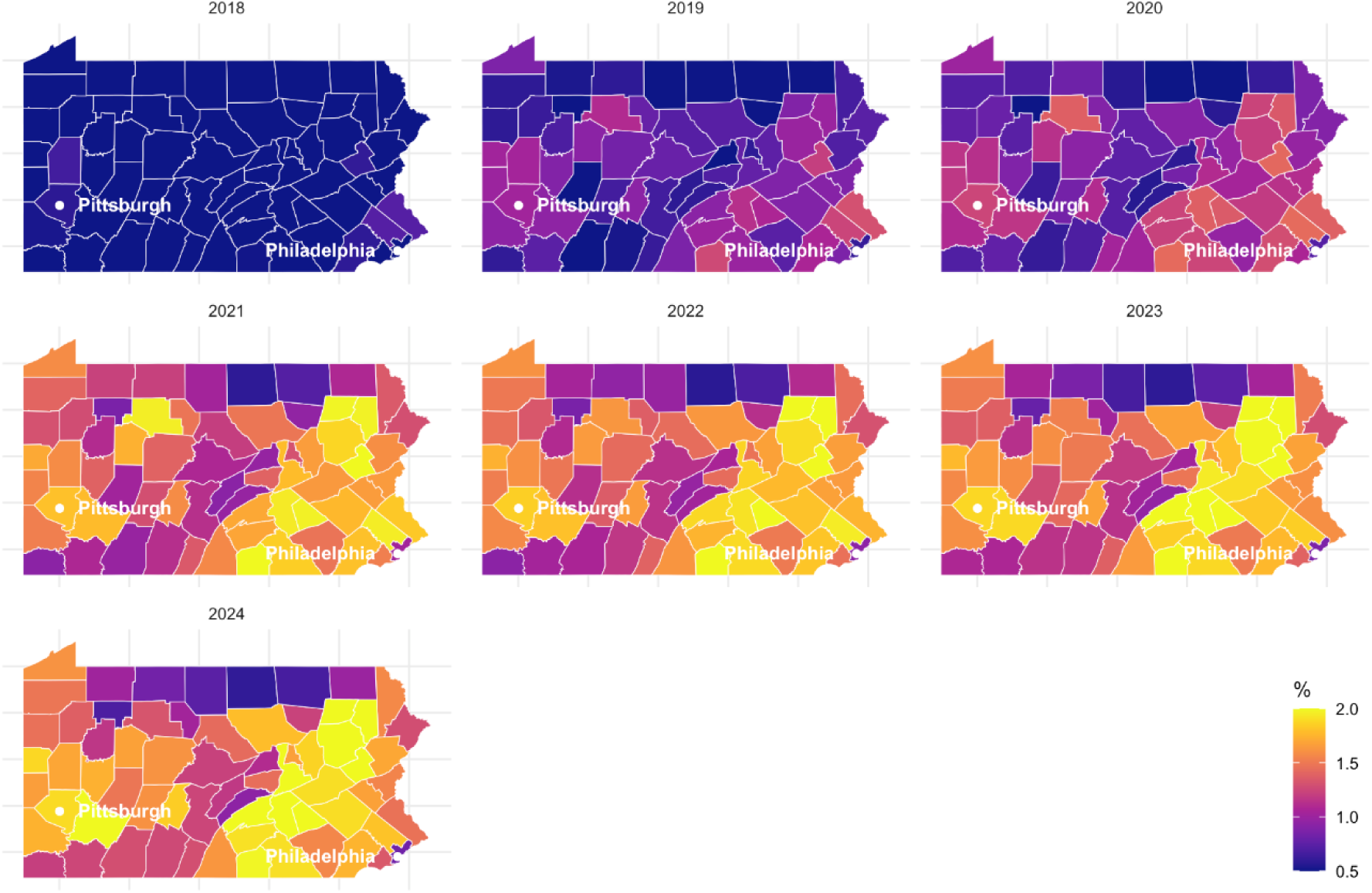
Heat map of the percent of adults in Pennsylvania with a medical cannabis certification for pain from 2018–2024 by county using proportional allocation of residential ratios.

**Supplementary Table 1.**
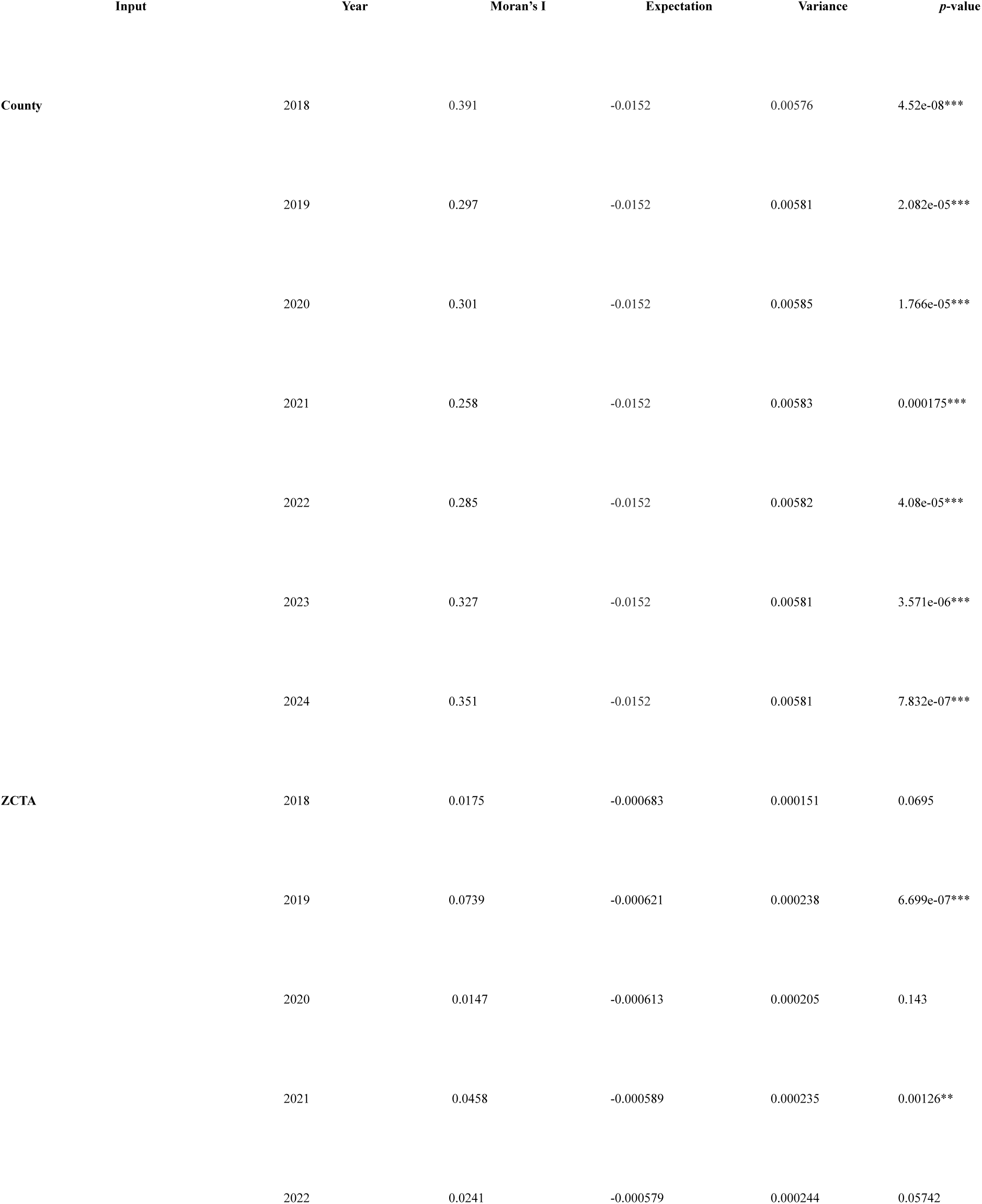
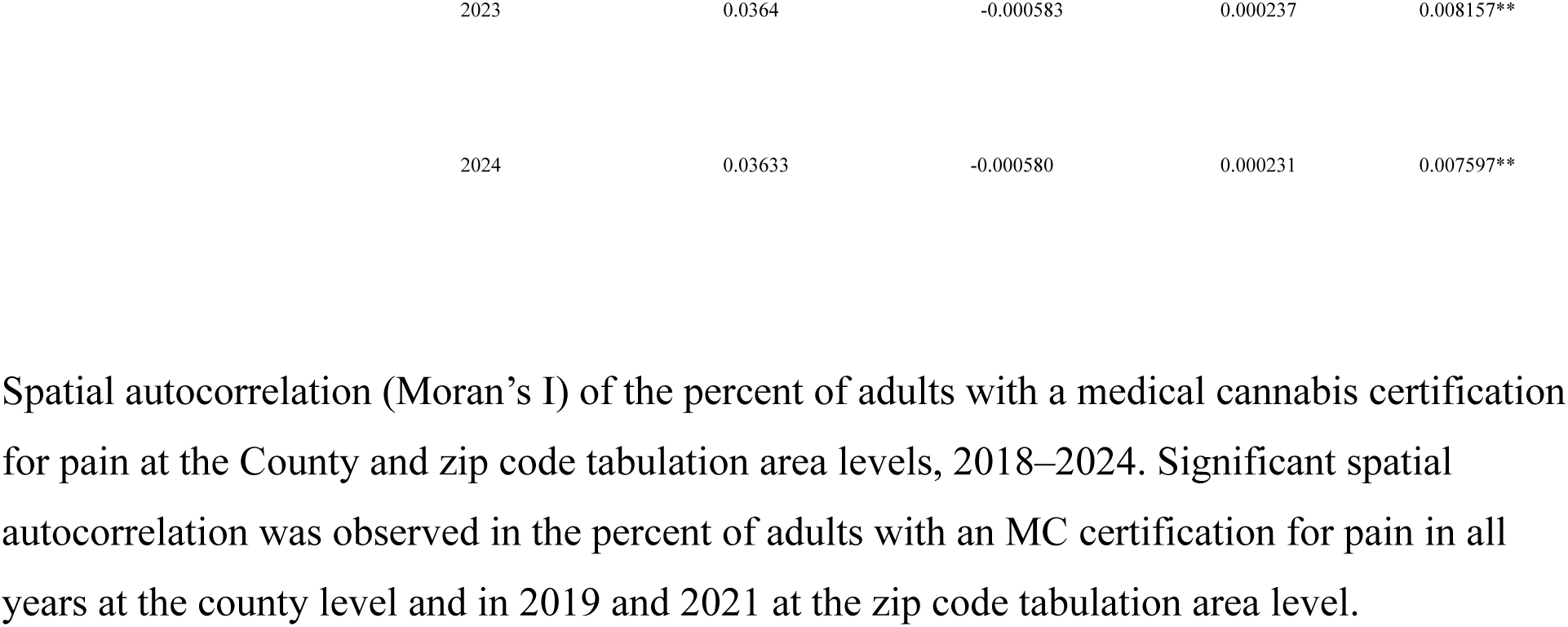
Spatial autocorrelation (Moran’s I) of the percent of adults with a medical cannabis certification for pain at the County and zip code tabulation area levels, 2018–2024. Significant spatial autocorrelation was observed in the percent of adults with an MC certification for pain in all years at the county level and in 2019 and 2021 at the zip code tabulation area level.

**Supplementary Fig. 4.**
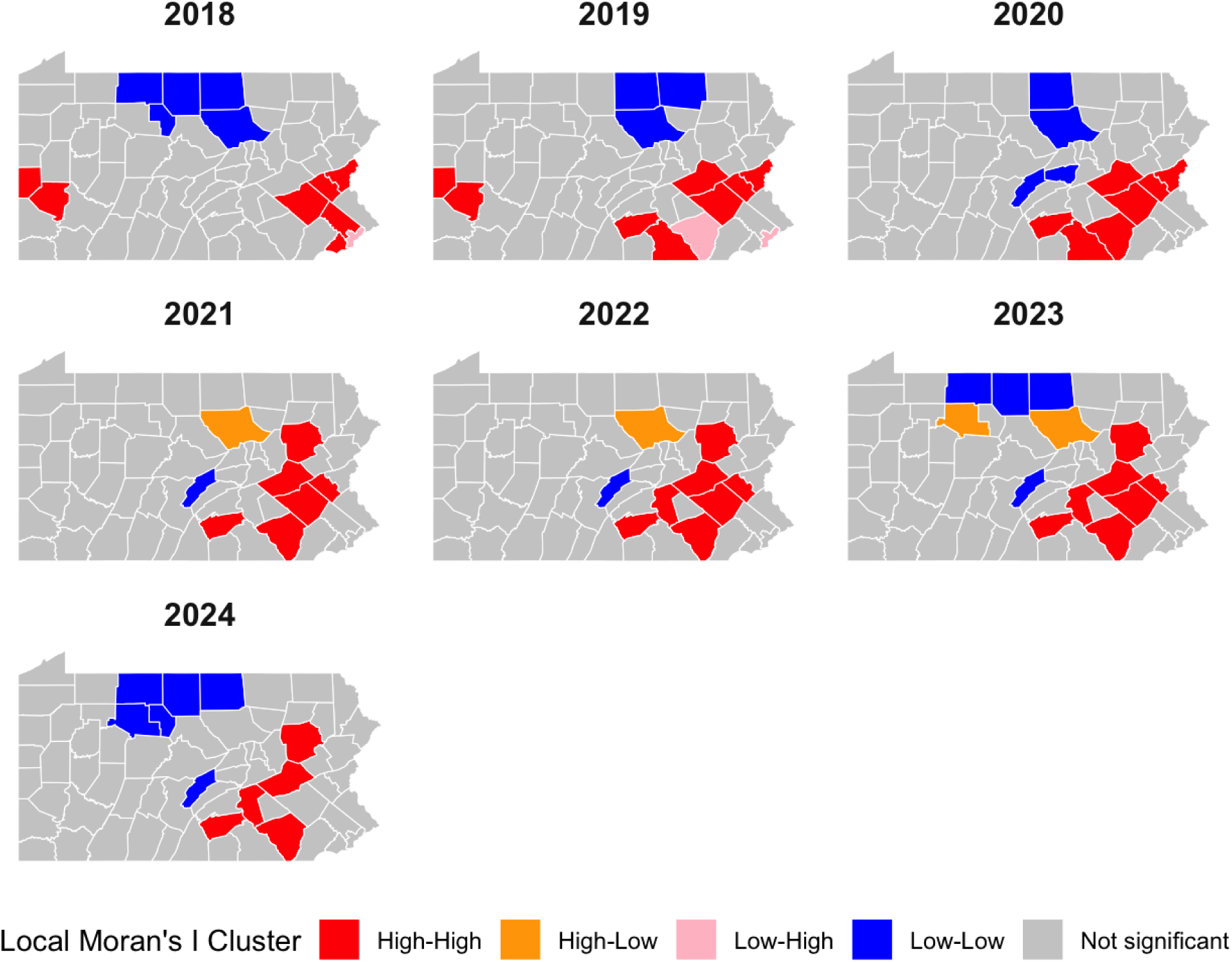
Pennsylvania map of local Moran’s I clusters from 2018 to 2024 at the county level without correction for multiple testing.

**Supplementary Fig. 5.**
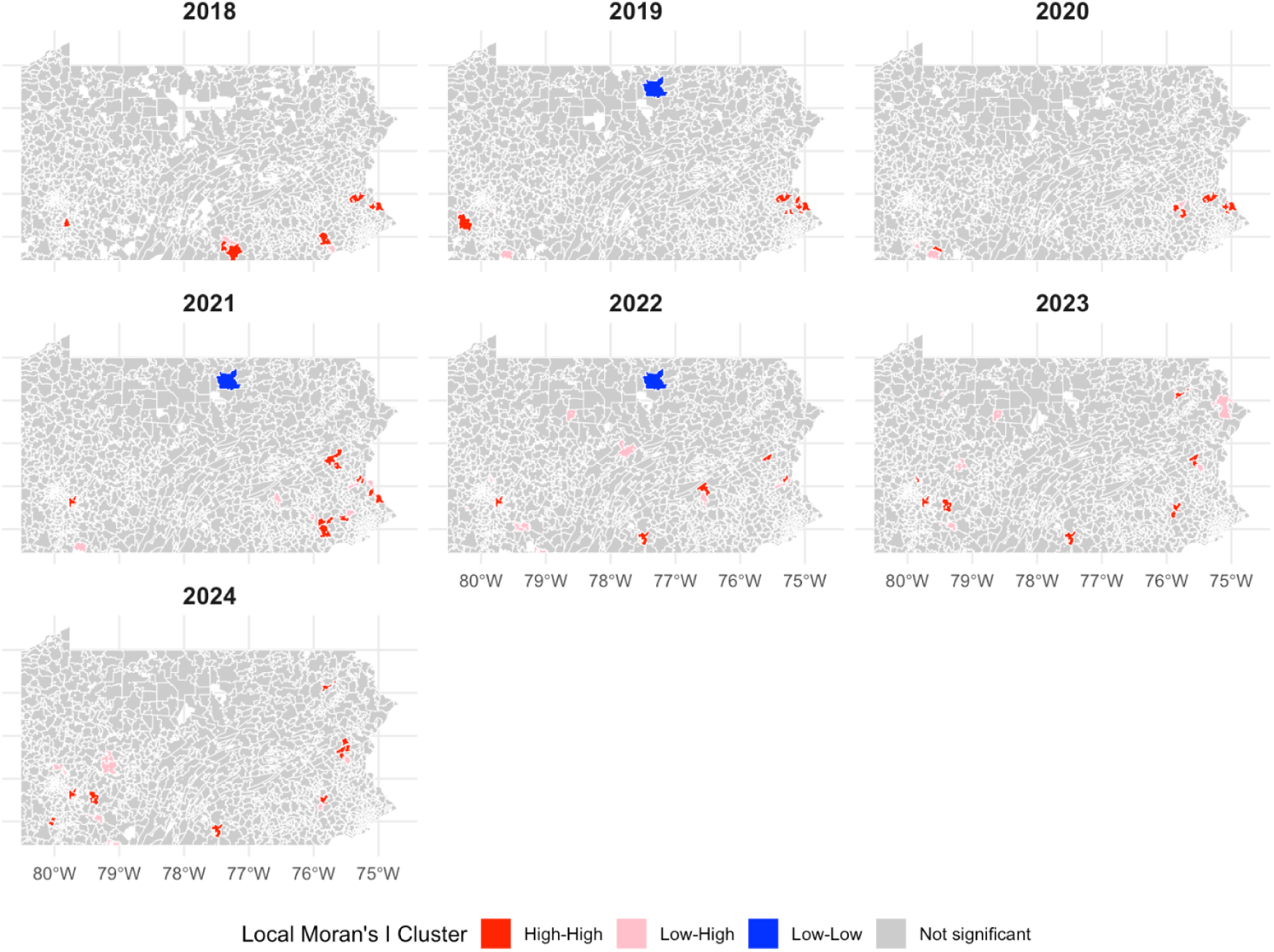
Pennsylvania map of local Moran’s I clusters of the percent of adults with a medical cannabis certification from pain from 2018 to 2024 at the zip code tabulation area level without correction for multiple testing.

**Supplementary Fig. 6.**
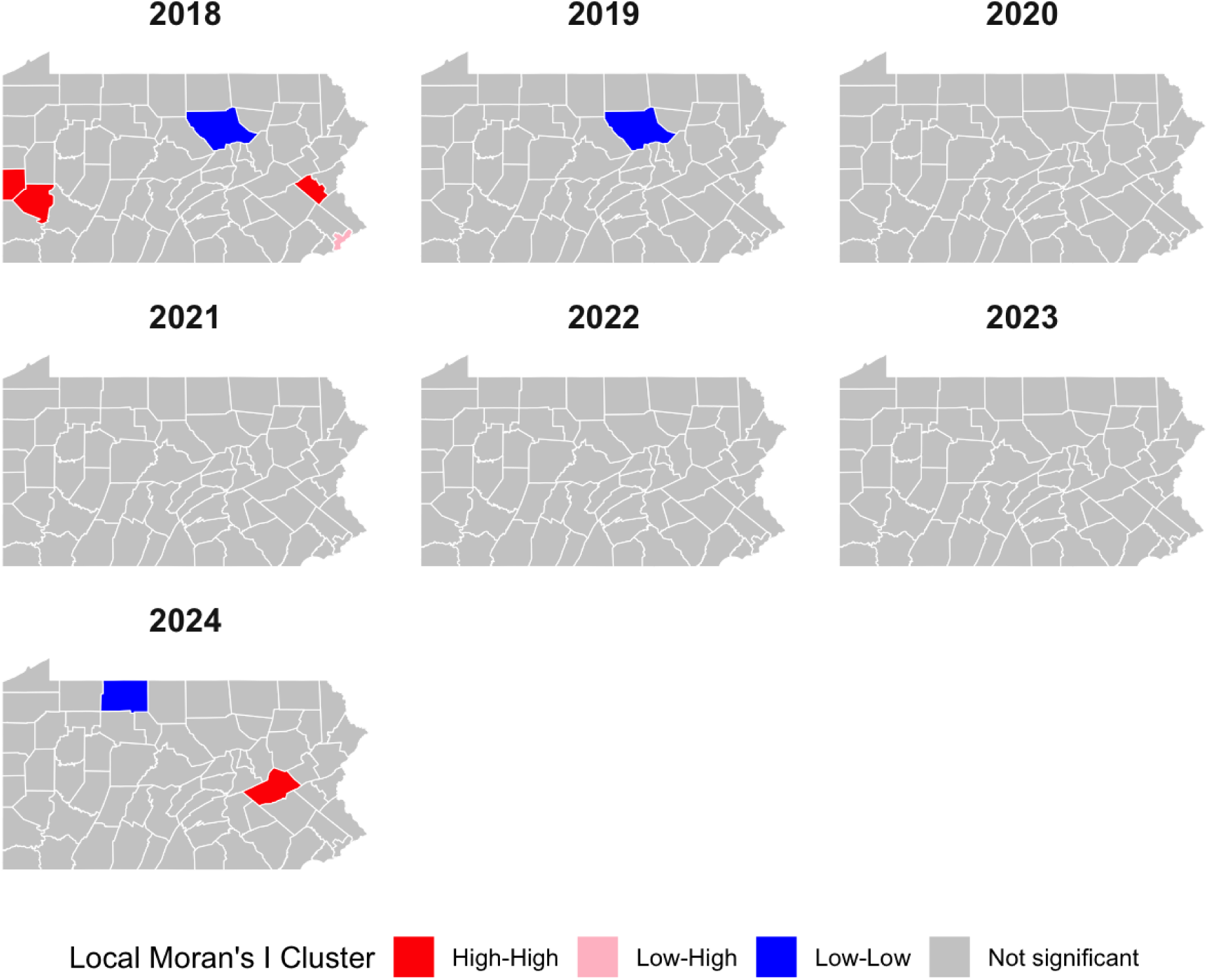
Pennsylvania map of local Moran’s I clusters of the percent of adults with a medical cannabis certification from pain from 2018 to 2024 corrected using the false discovery rate. Lycoming County was a coldspot in 2018 and 2019 and McKean County was a coldspot in 2024. Beaver County, Allegheny County, and Lehigh County were hotspots in 2018 and Schuylkill County was a hotspot in 2024. Philadelphia County was a low-high spatial outlier in 2018.

**Supplementary Fig. 7.**
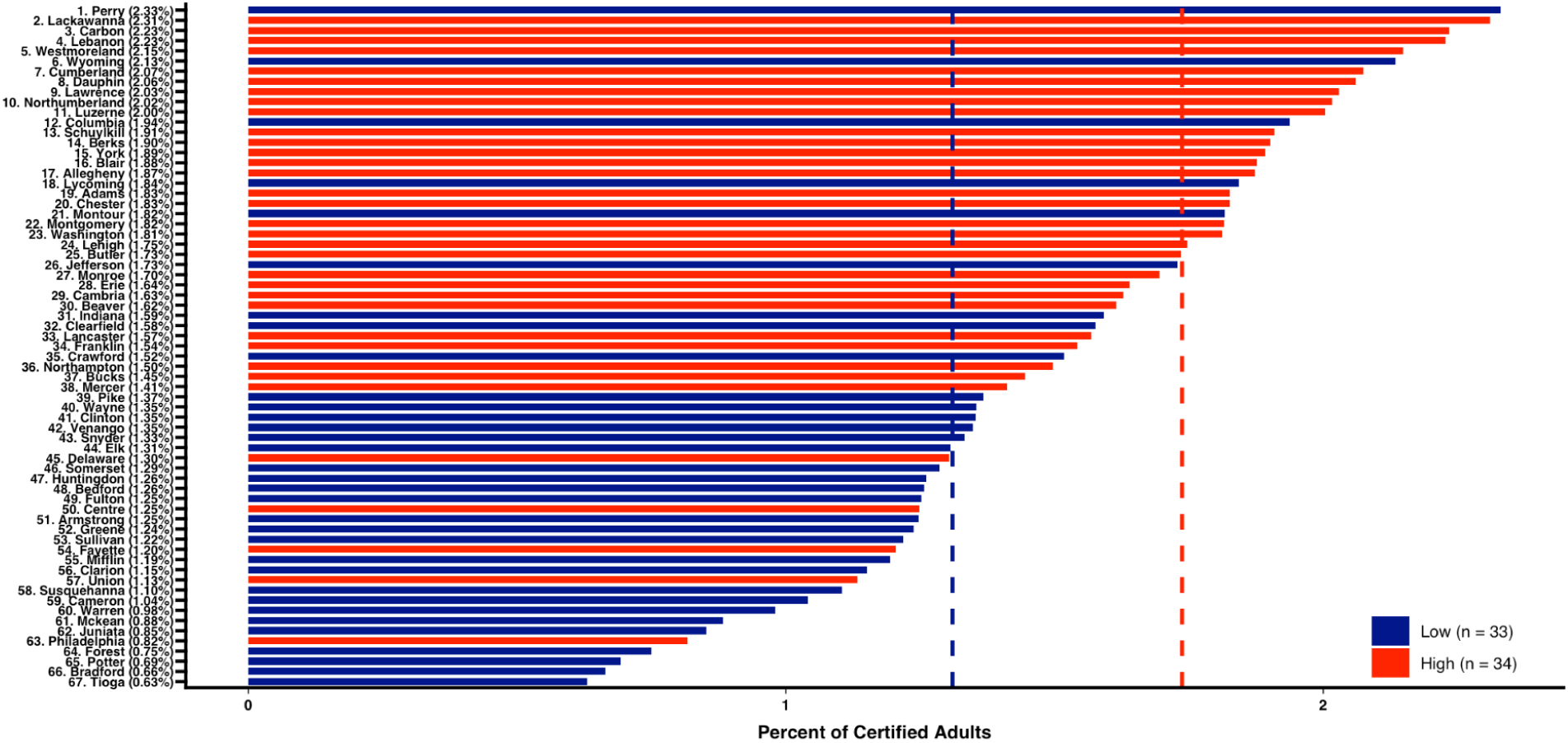
Waterfall plot of the percent of adults certified for medical cannabis for pain in Pennsylvania by county and by population density of adults. There was a significantly higher proportion of MC certifications for pain in counties with higher population densities of adults (1.76 +/− 0.12%) than counties with smaller (1.38% +/− 0.14%) population densities (t(65) = 4.66, p < 0.001, *d* = 1.14).

**Supplementary Fig. 8.**
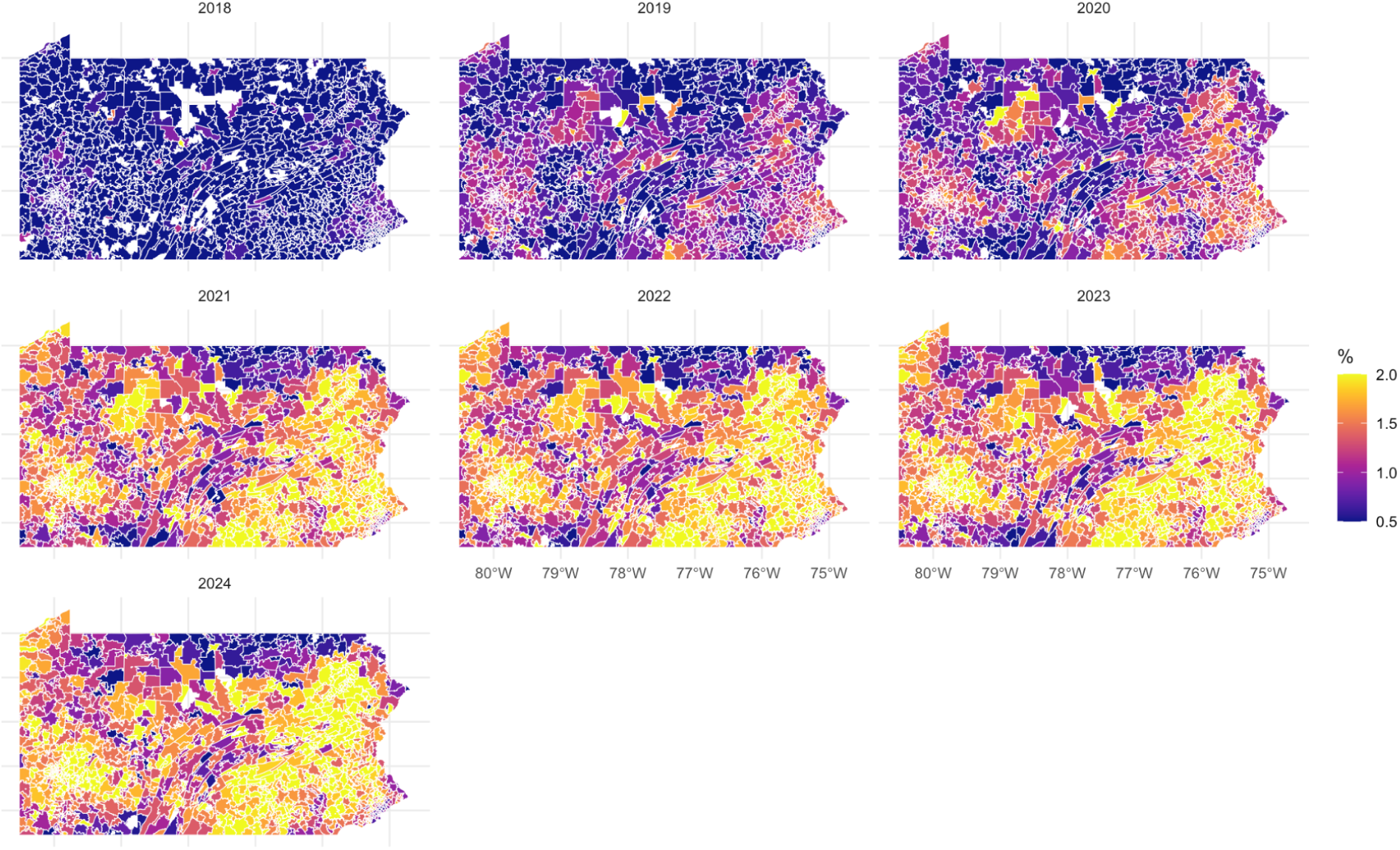
Heat map of the percent of adults in Pennsylvania with a Medical Cannabis certification for pain from 2018–2024 at the Zip Code Tabulation Area (ZCTA) level. Blank spaces indicate ZCTAs with no certifications.

**Supplementary Fig. 9.**
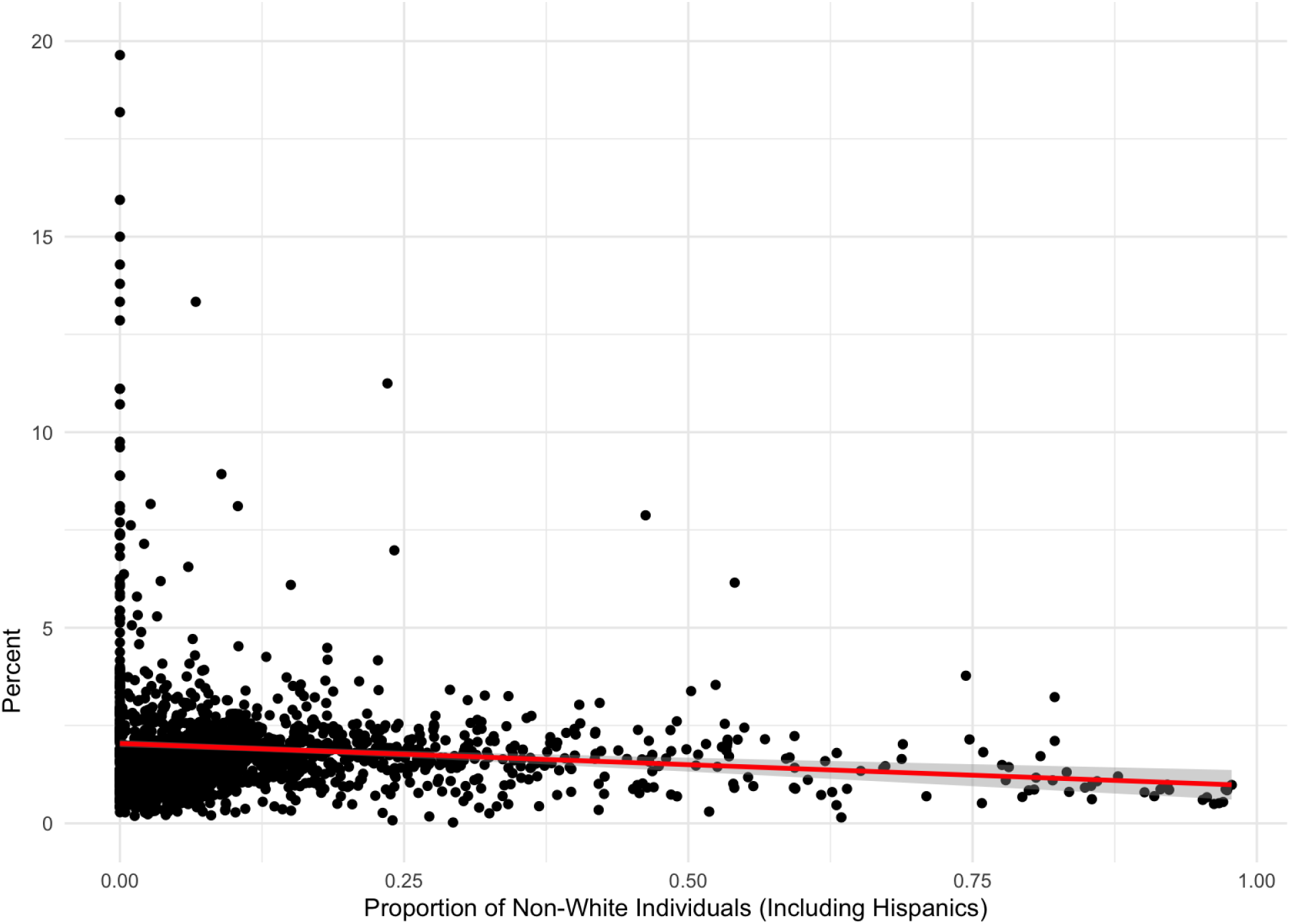
Scatterplot of the percent of adults with a medical cannabis certification for pain in Pennsylvania versus the proportion of non-White individuals, including Hispanics, in 2024 at the ZCTA level (r(1,722) = –0.0749, p = 0.001885)

**Supplementary Fig. 10.**
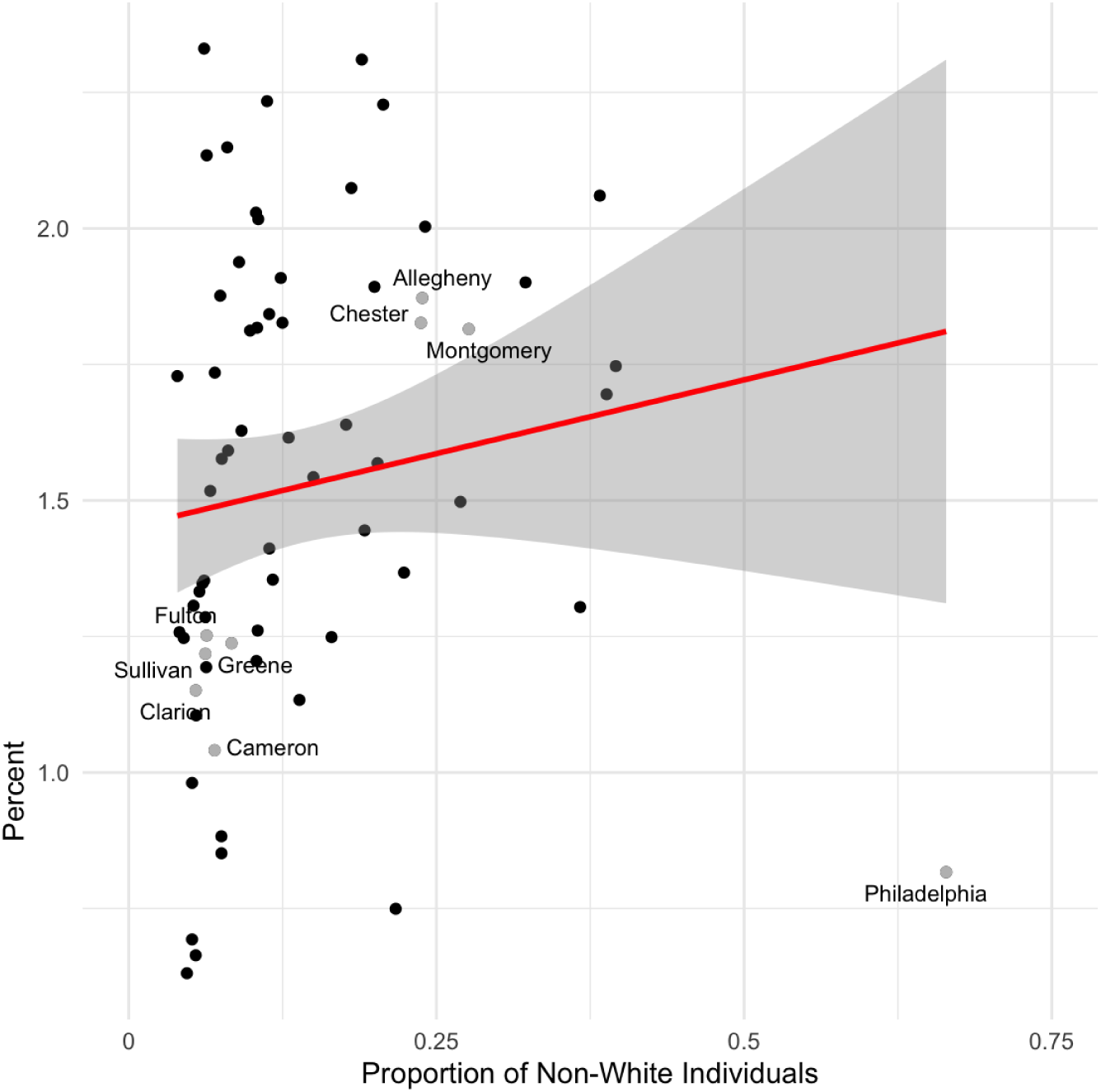
Scatterplot of the percent of adults with a medical cannabis certification for pain versus the proportion of non-White individuals, including Hispanics, in 2024 at the county level (r(65) = 0.142, p = 0.2505)

**Supplementary Fig. 11.**
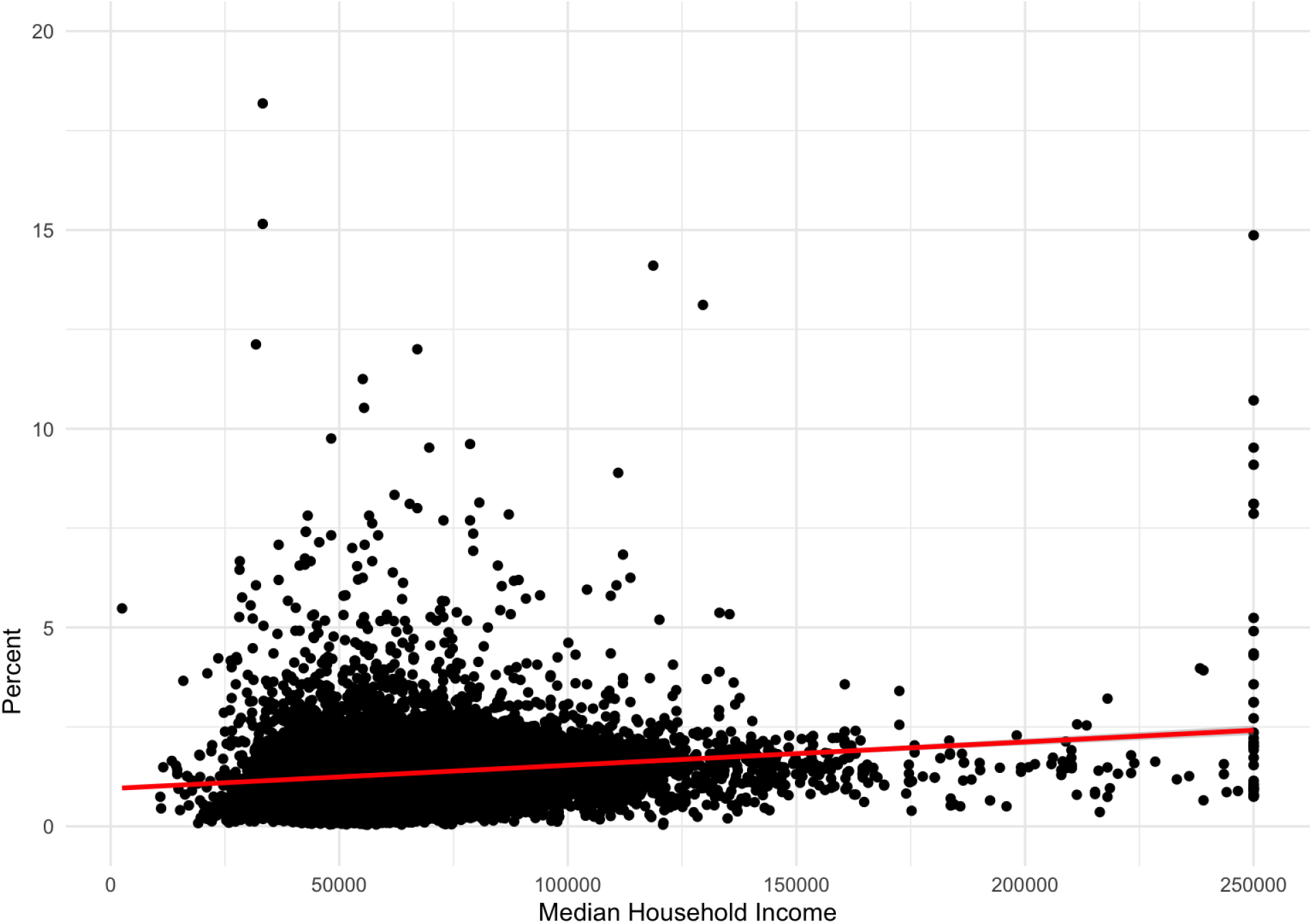
Scatterplot of the percent of adults with a medical cannabis certification for pain versus median household income in 2024 at the ZCTA level (r(1,606) = 0.0278, p = 0.264)

**Supplementary Fig. 12.**
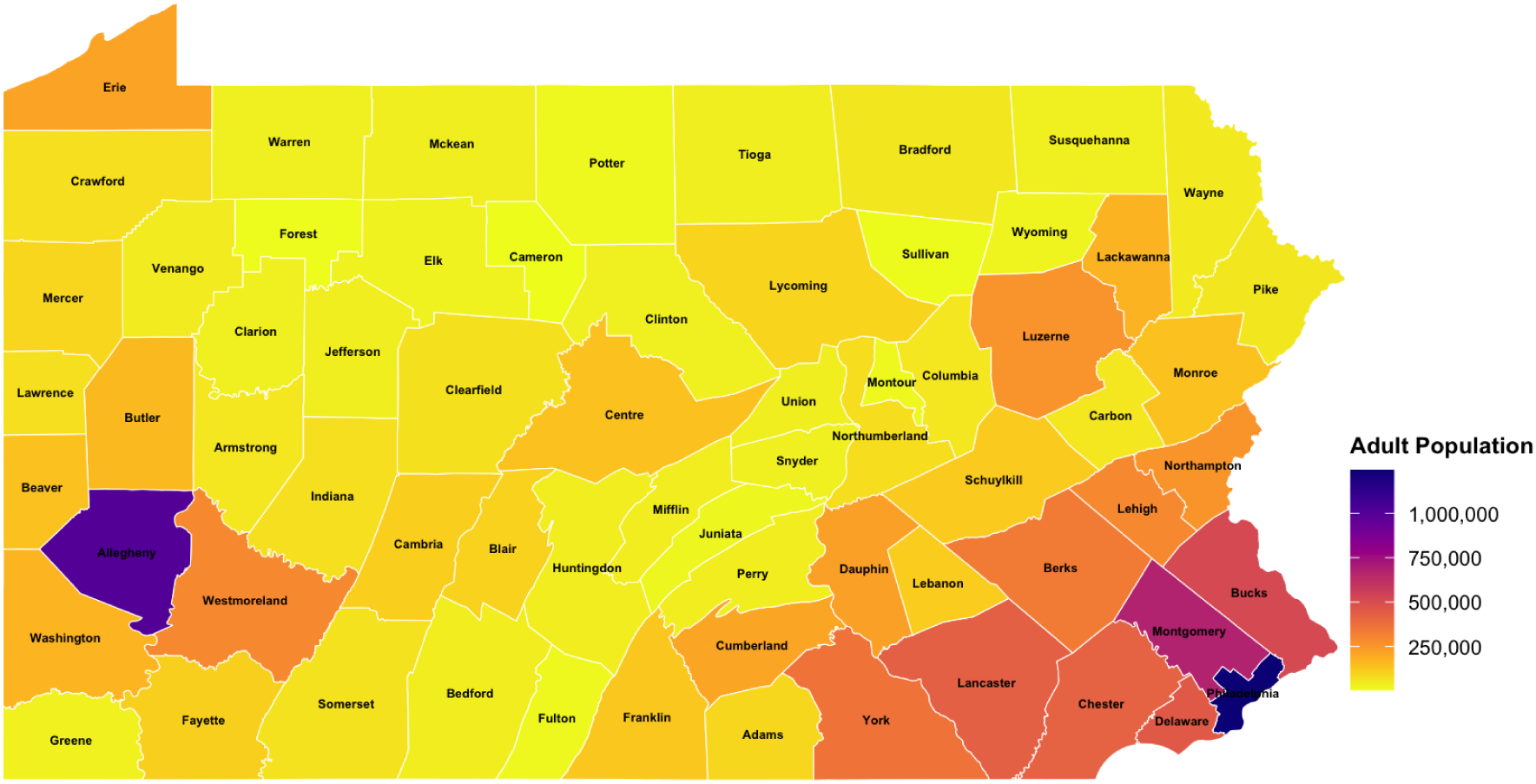
Heat map of the adult population in the counties in Pennsylvania. Over half (55.5%) of adults in Pennsylvania in 2024 resided in ten counties (Philadelphia, Allegheny, Montgomery, Bucks, Delaware, Lancaster, Chester, York, Berks, Lehigh).

**Supplementary Fig. 13.**
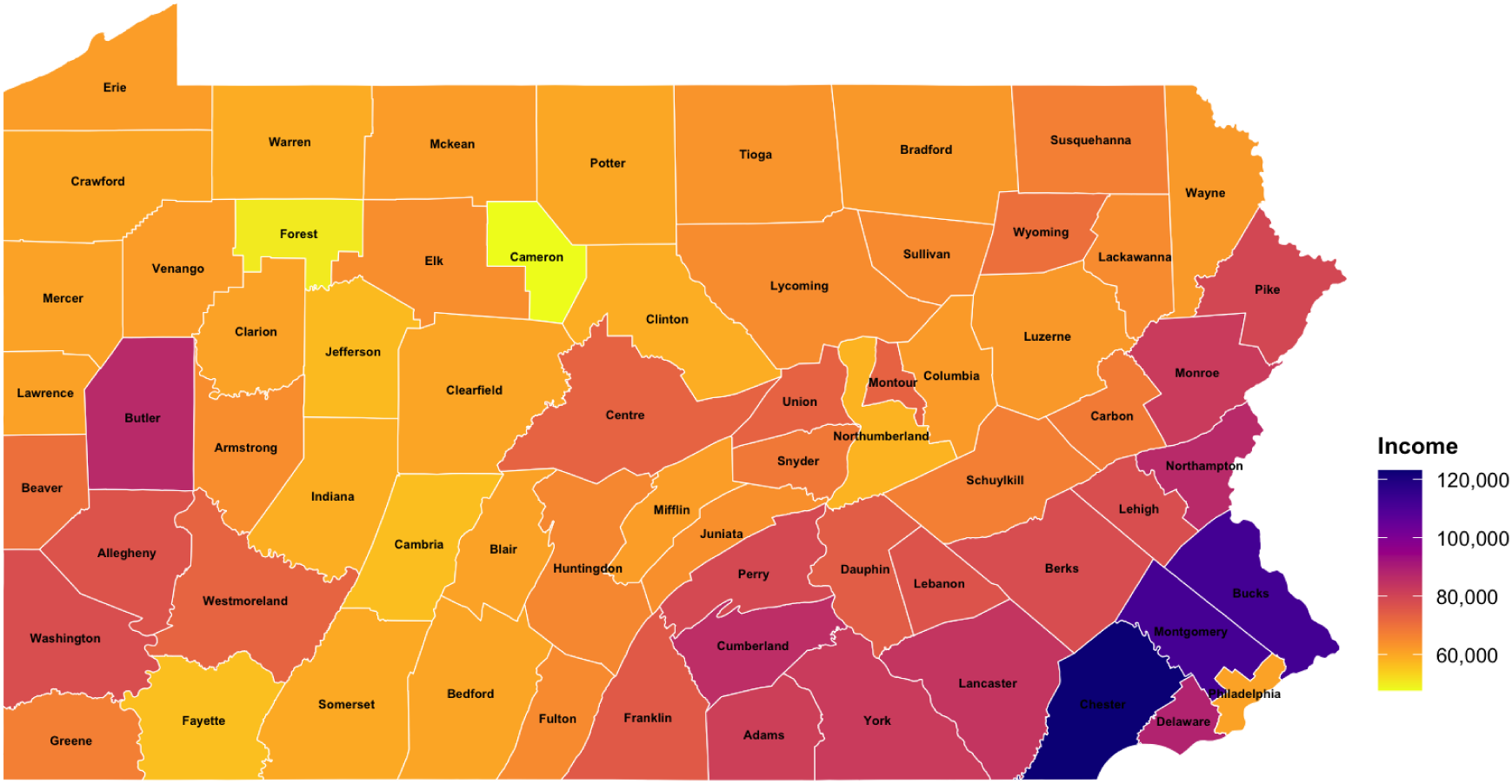
Heat map of the median household income in the counties in Pennsylvania in 2024. Pittsburgh is in Allegheny County while Philadelphia is in Philadelphia County.

