## Supplementary Material for "Medical Cannabis Certifications for Severe Chronic and Intractable Pain: Discerning Geographic Patterns Across Pennsylvania, USA"

**Supplementary Fig. 1.**

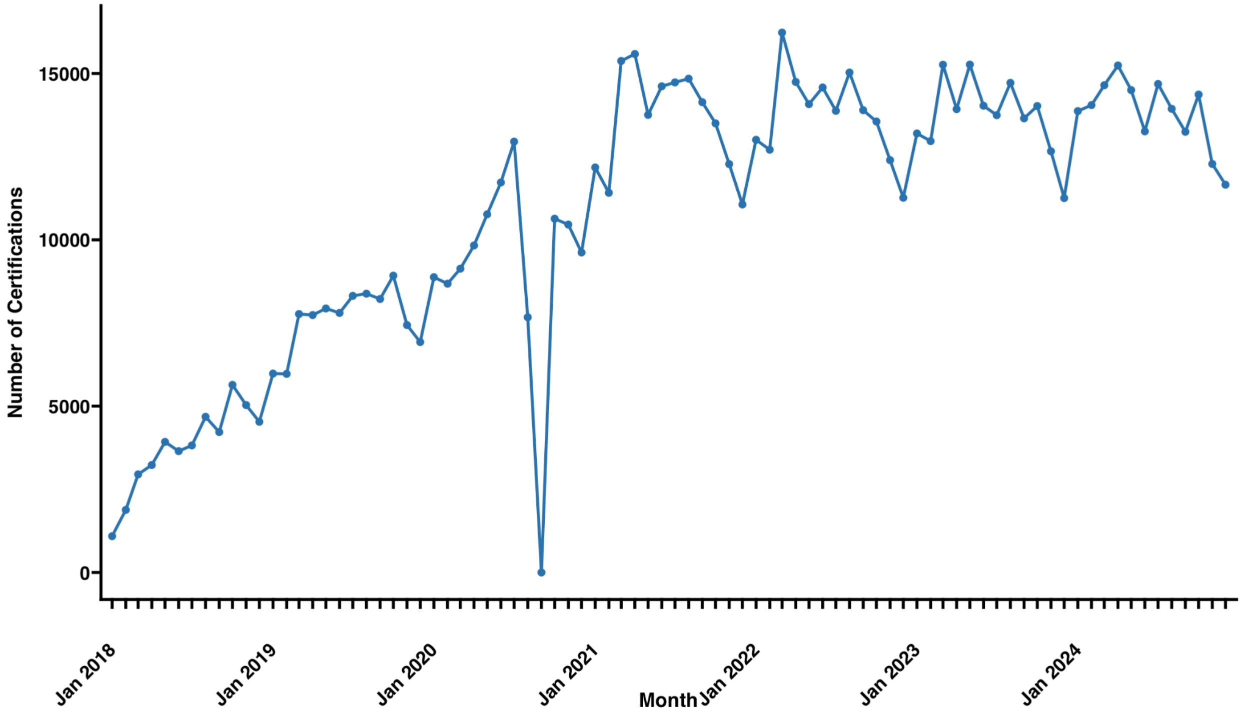


Monthly counts of MC certifications for pain. No MC certifications for pain were reported in September 2020.

**Supplementary Fig. 2.**

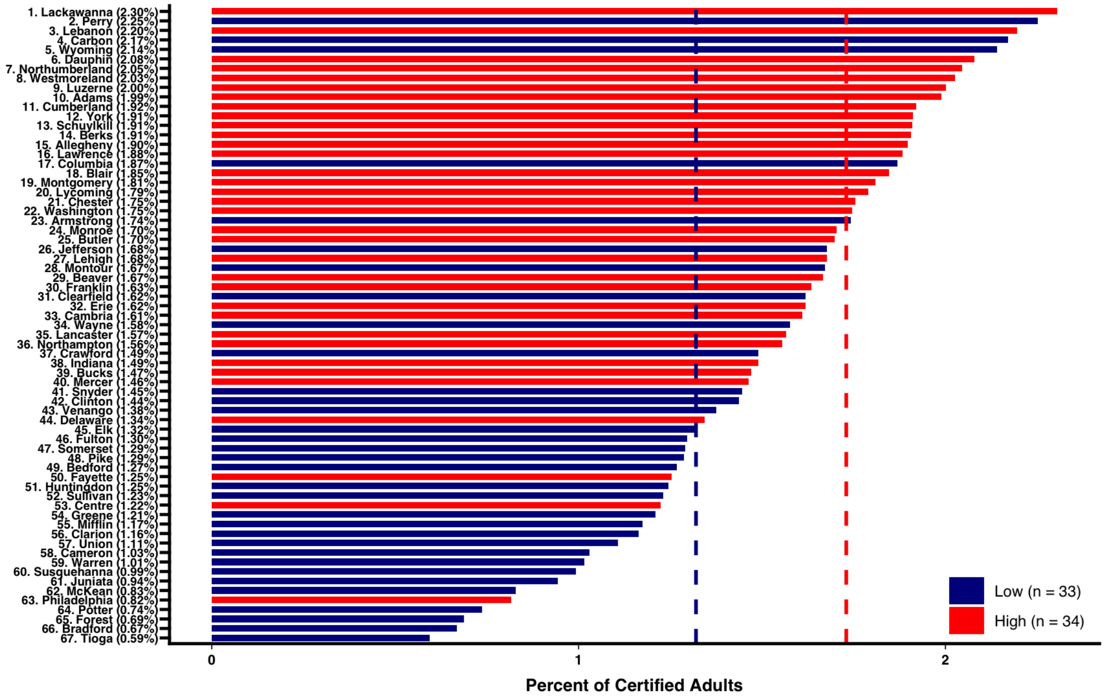


Waterfall plot of the percent of adults certified for medical cannabis for pain in Pennsylvania by county and by population size using proportional allocation of residential ratios. Using proportional allocation, there was a significantly higher proportion of MC certifications for pain in more populated (1.73 +/- 0.10%) than less populated (1.32% +/- 0.15%) counties (t(65) = 4.60, p < 0.001, *d* = 1.22).

**Supplementary Fig. 3.**

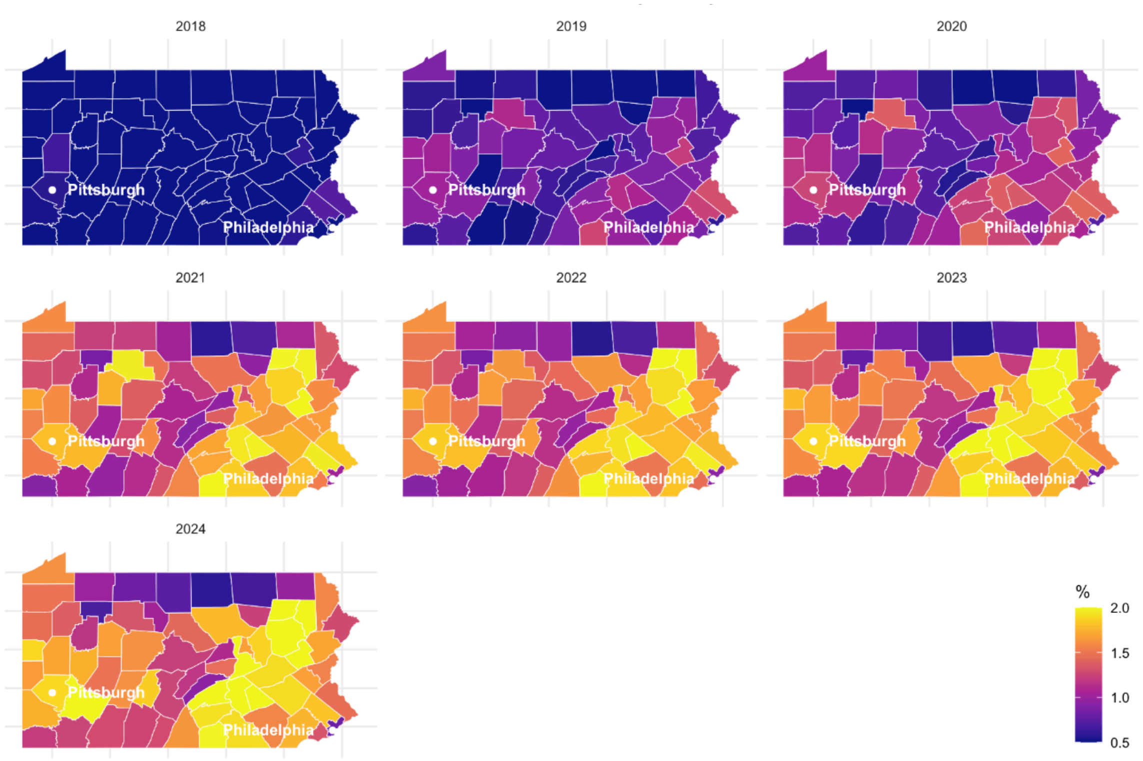


Heat map of the percent of adults in Pennsylvania with a medical cannabis certification for pain from 2018–2024 by county using proportional allocation of residential ratios

### Supplementary Table 1.

| Input | Year | Moran's I | Expectation | Variance | *p*-value |
|---|---|---|---|---|---|
| **County** | 2018 | 0.391 | -0.0152 | 0.00576 | 4.52e-08*** |
| | 2019 | 0.297 | -0.0152 | 0.00581 | 2.082e-05*** |
| | 2020 | 0.301 | -0.0152 | 0.00585 | 1.766e-05*** |
| | 2021 | 0.258 | -0.0152 | 0.00583 | 0.000175*** |
| | 2022 | 0.285 | -0.0152 | 0.00582 | 4.08e-05*** |
| | 2023 | 0.327 | -0.0152 | 0.00581 | 3.571e-06*** |
| | 2024 | 0.351 | -0.0152 | 0.00581 | 7.832e-07*** |
| **ZCTA** | 2018 | 0.0175 | -0.000683 | 0.000151 | 0.0695 |
| | 2019 | 0.0739 | -0.000621 | 0.000238 | 6.699e-07*** |
| | 2020 | 0.0147 | -0.000613 | 0.000205 | 0.143 |
| | 2021 | 0.0458 | -0.000589 | 0.000235 | 0.00126** |
| | 2022 | 0.0241 | -0.000579 | 0.000244 | 0.05742 |

| | | | | |
|---|---|---|---|---|
| 2023 | 0.0364 | -0.000583 | 0.000237 | 0.008157** |
| 2024 | 0.03633 | -0.000580 | 0.000231 | 0.007597** |

Spatial autocorrelation (Moran's I) of the percent of adults with a medical cannabis certification for pain at the County and zip code tabulation area levels, 2018–2024. Significant spatial autocorrelation was observed in the percent of adults with an MC certification for pain in all years at the county level and in 2019 and 2021 at the zip code tabulation area level.

**Supplementary Fig. 4.**

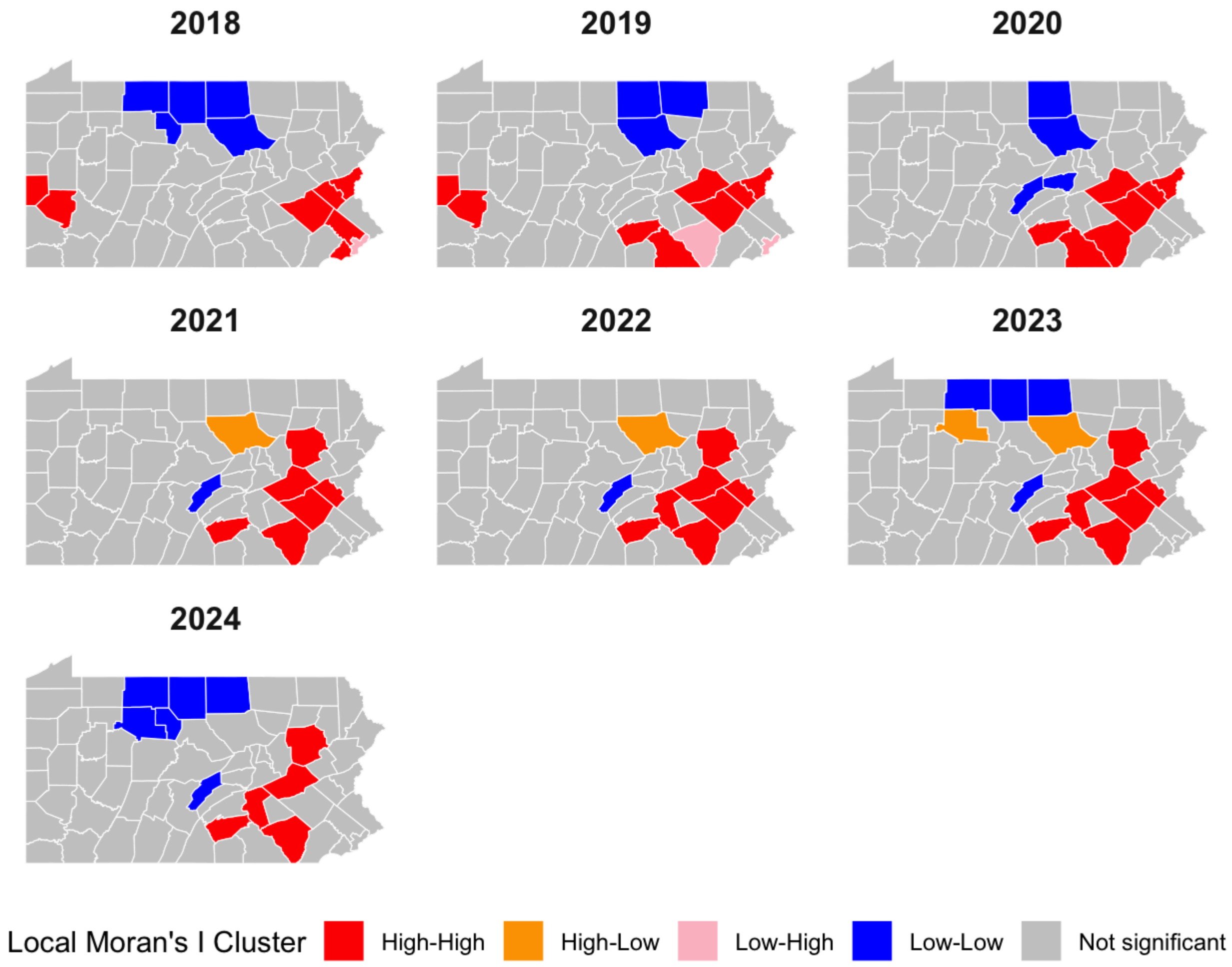


Pennsylvania map of local Moran's I clusters from 2018 to 2024 at the county level without correction for multiple testing

**Supplementary Fig. 5.**

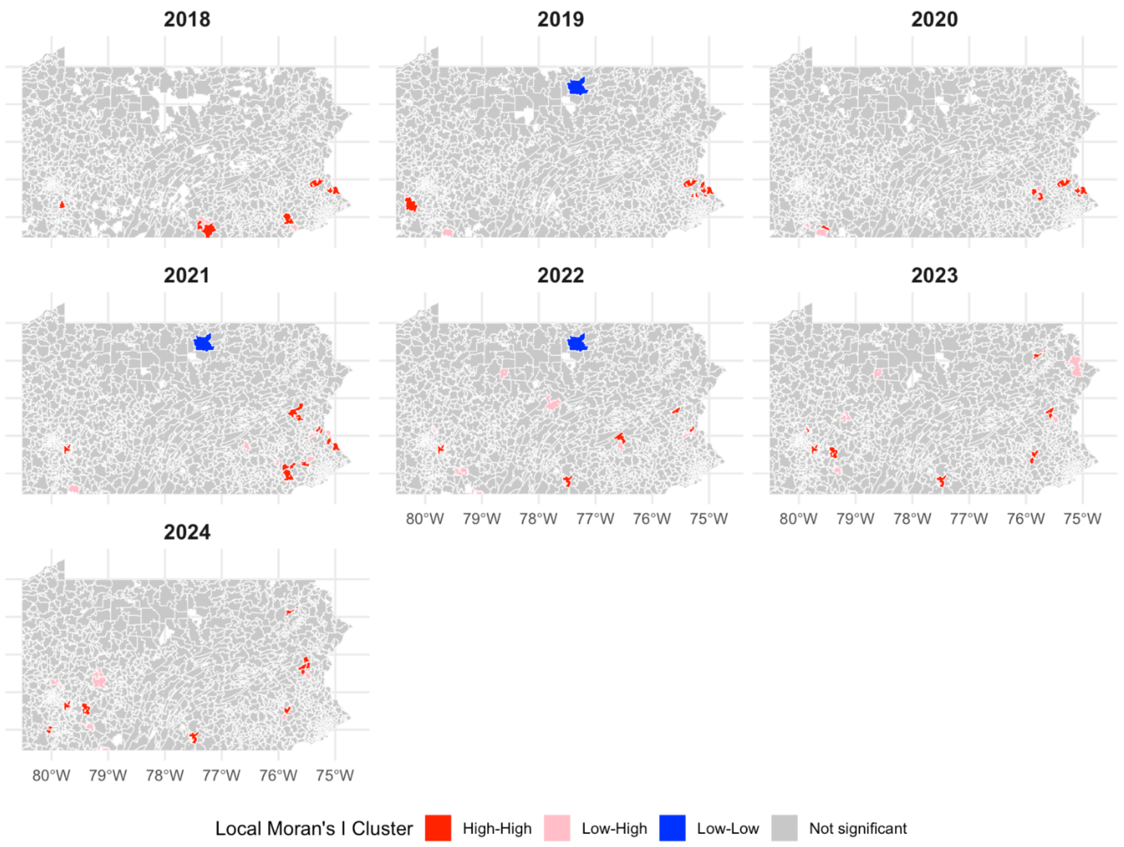


Pennsylvania map of local Moran’s I clusters of the percent of adults with a medical cannabis certification from pain from 2018 to 2024 at the zip code tabulation area level without correction for multiple testing

**Supplementary Fig. 6.**

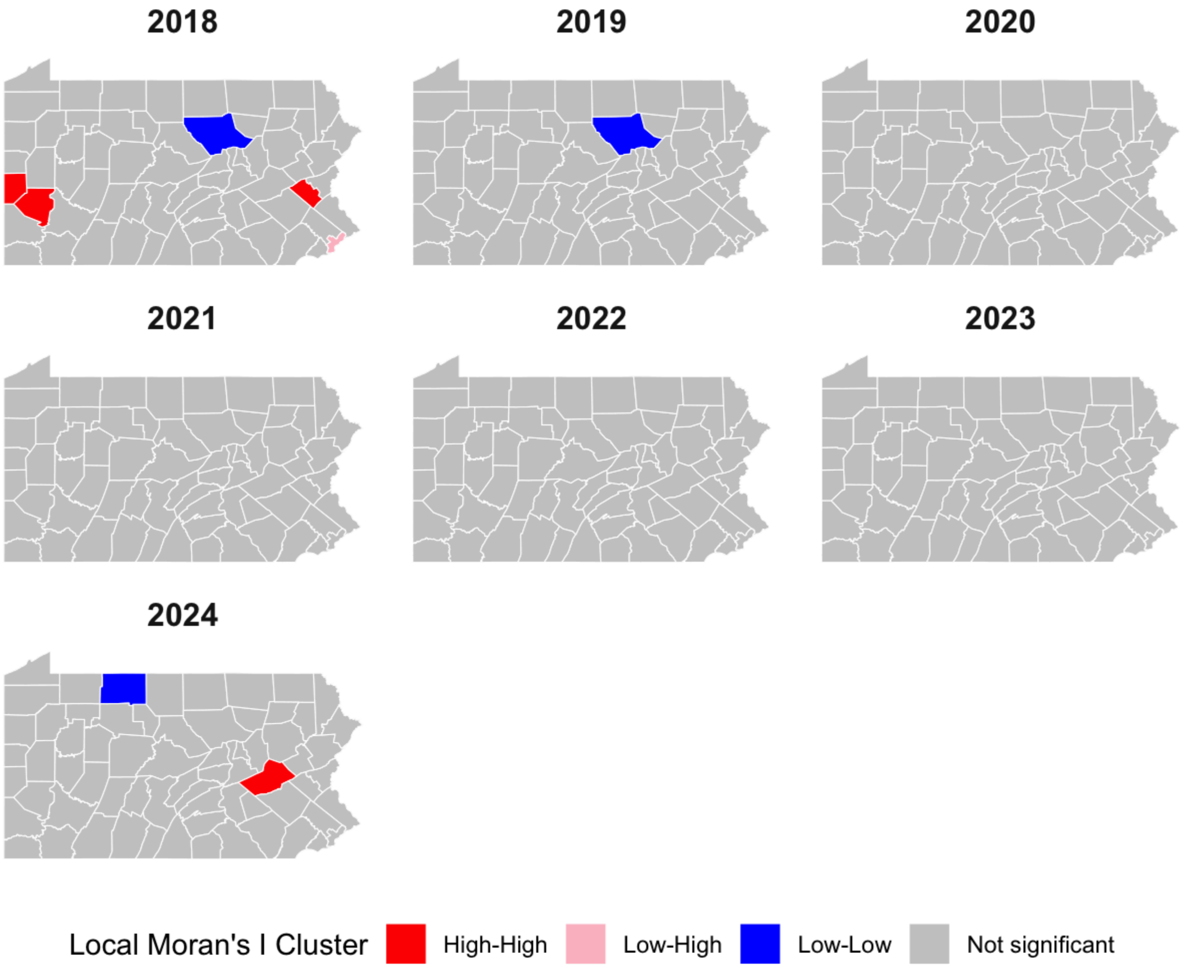


Pennsylvania map of  local Moran's I clusters of the percent of adults with a medical cannabis certification from pain from 2018 to 2024 corrected using the false discovery rate. Lycoming County was a coldspot in 2018 and 2019 and McKean County was a coldspot in 2024. Beaver County, Allegheny County, and Lehigh County were hotspots in 2018 and Schuylkill County was a hotspot in 2024. Philadelphia County was a low-high spatial outlier in 2018.

**Supplementary Fig. 7.**

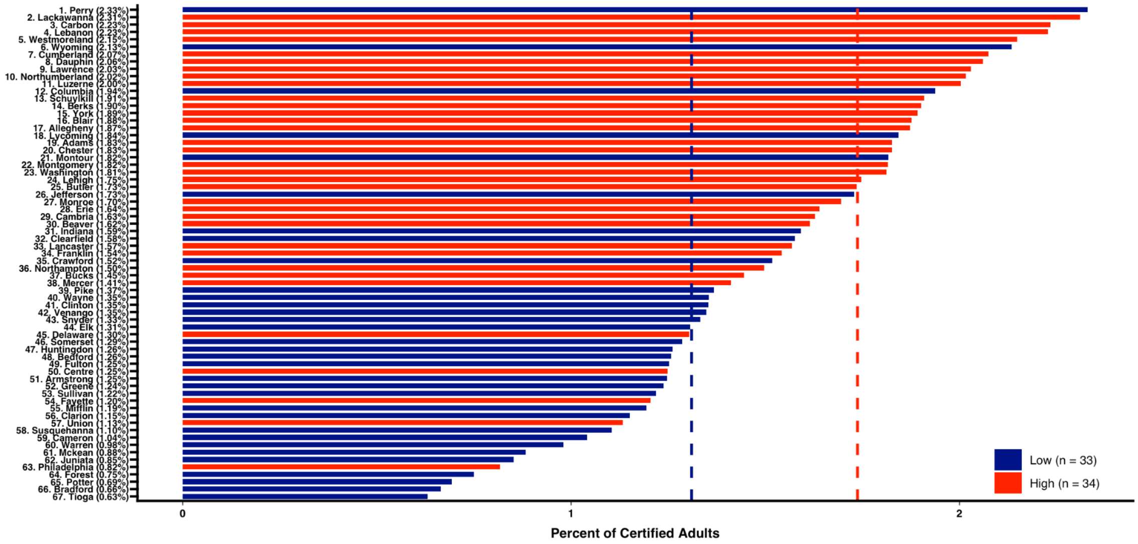


Waterfall plot of the percent of adults certified for medical cannabis for pain in Pennsylvania by county and by population density of adults. There was a significantly higher proportion of MC certifications for pain in counties with higher population densities of adults (1.76 +/- 0.12%) than counties with smaller (1.38% +/- 0.14%) population densities ($t(65) = 4.66$, $p < 0.001$, $d = 1.14$).

**Supplementary Fig. 8.**

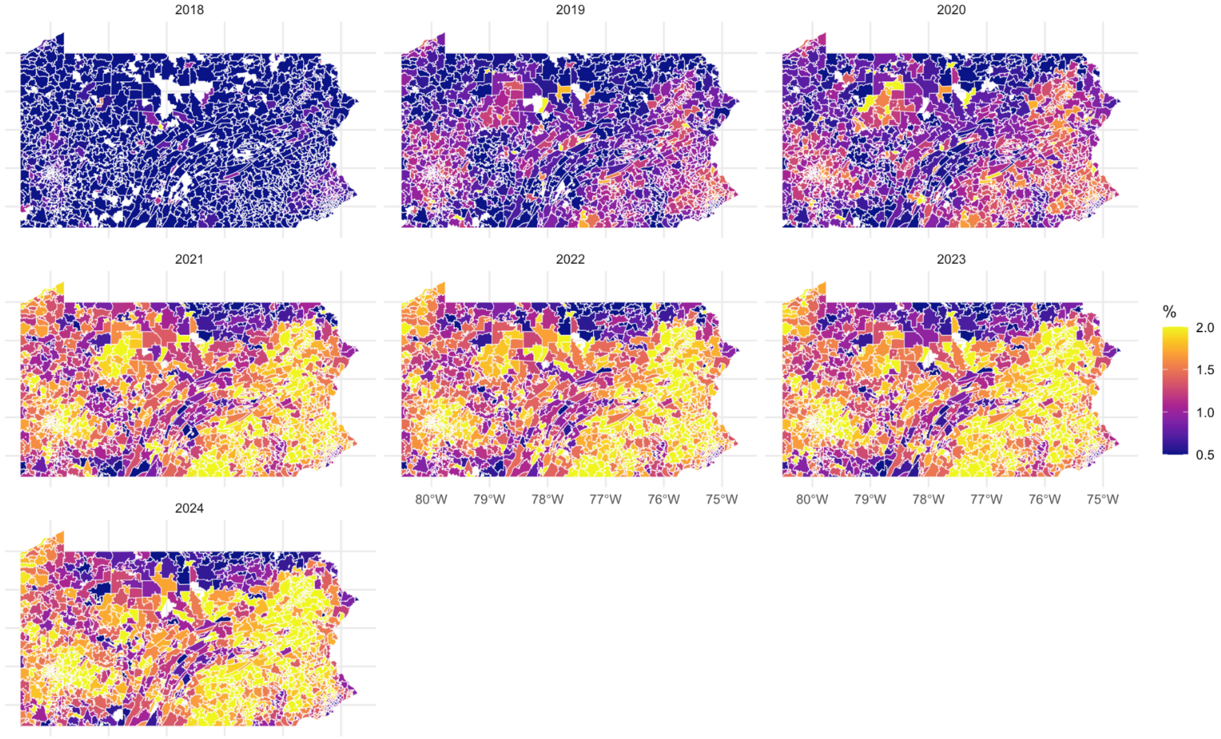


Heat map of the percent of adults in Pennsylvania with a Medical Cannabis certification for pain from 2018–2024 at the Zip Code Tabulation Area (ZCTA) level. Blank spaces indicate ZCTAs with no certifications.

**Supplementary Fig. 9.**

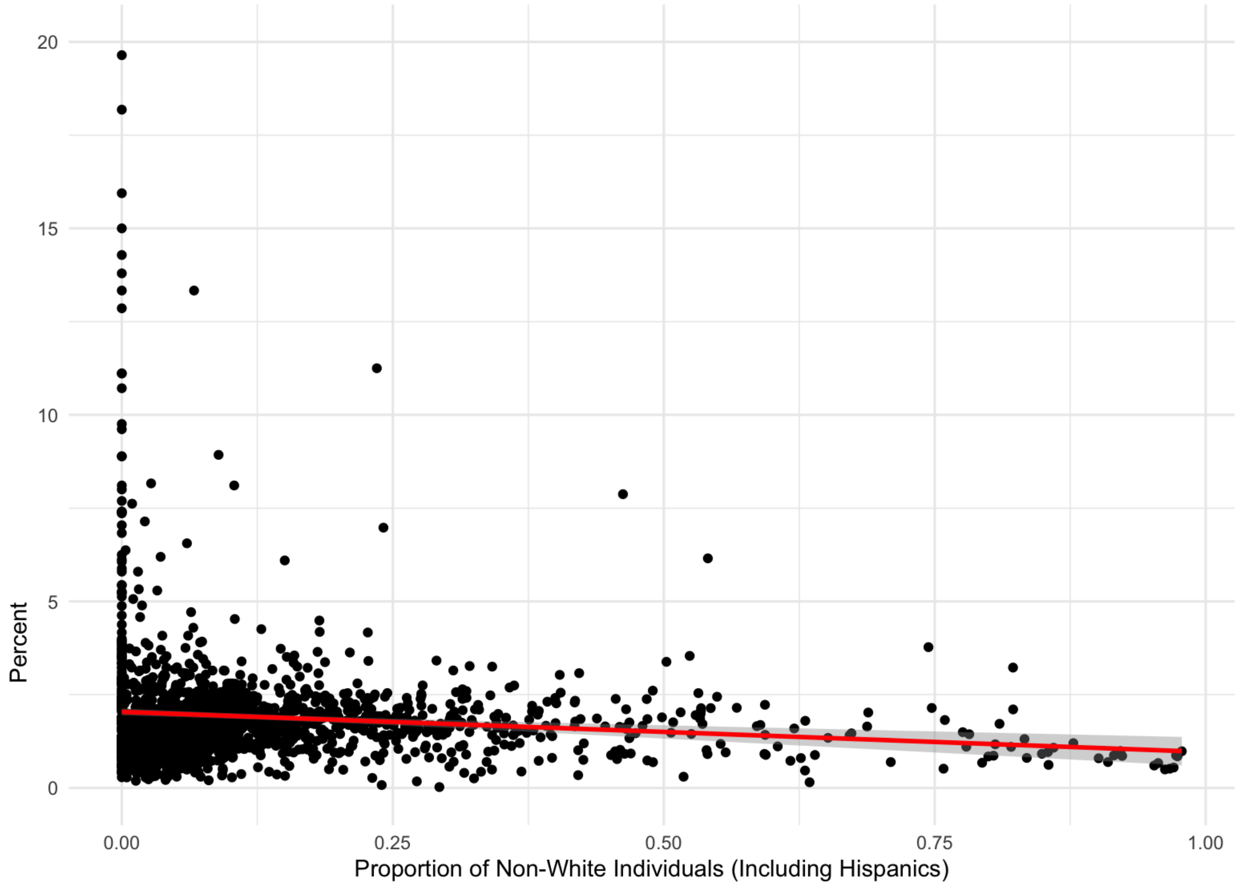


Scatterplot of the percent of adults with a medical cannabis certification for pain in Pennsylvania versus the proportion of non-White individuals, including Hispanics, in 2024 at the ZCTA level ($r(1,722) = -0.0749$, $p = 0.001885$)

**Supplementary Fig. 10.**

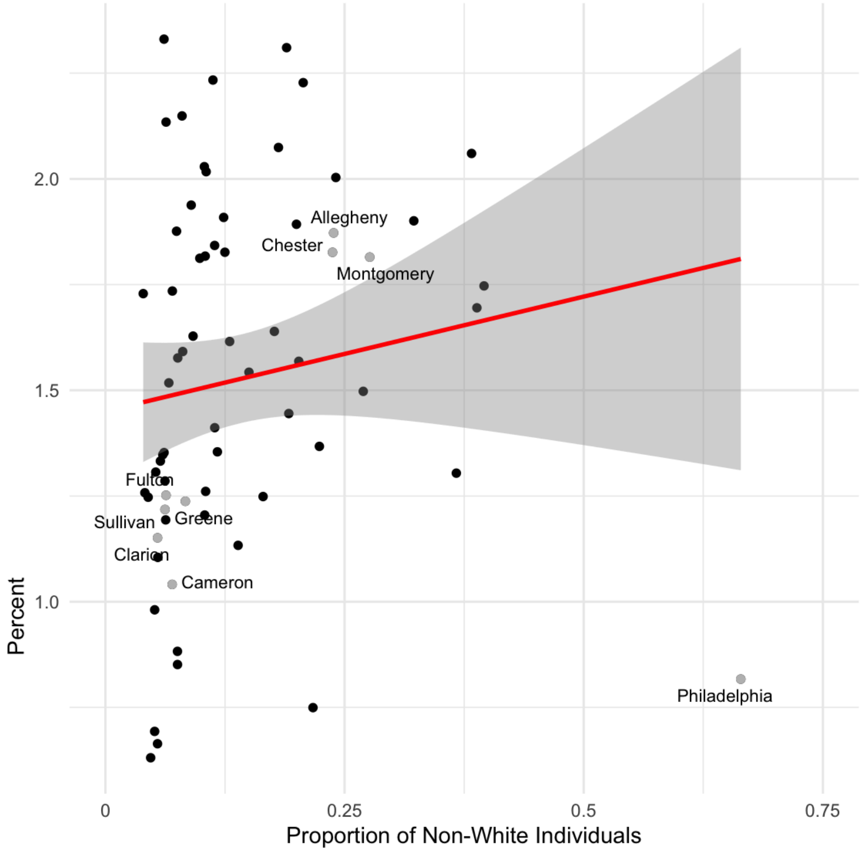


Scatterplot of the percent of adults with a medical cannabis certification for pain versus the proportion of non-White individuals, including Hispanics, in 2024 at the county level ($r(65) = 0.142$, $p = 0.2505$)

**Supplementary Fig. 11.**

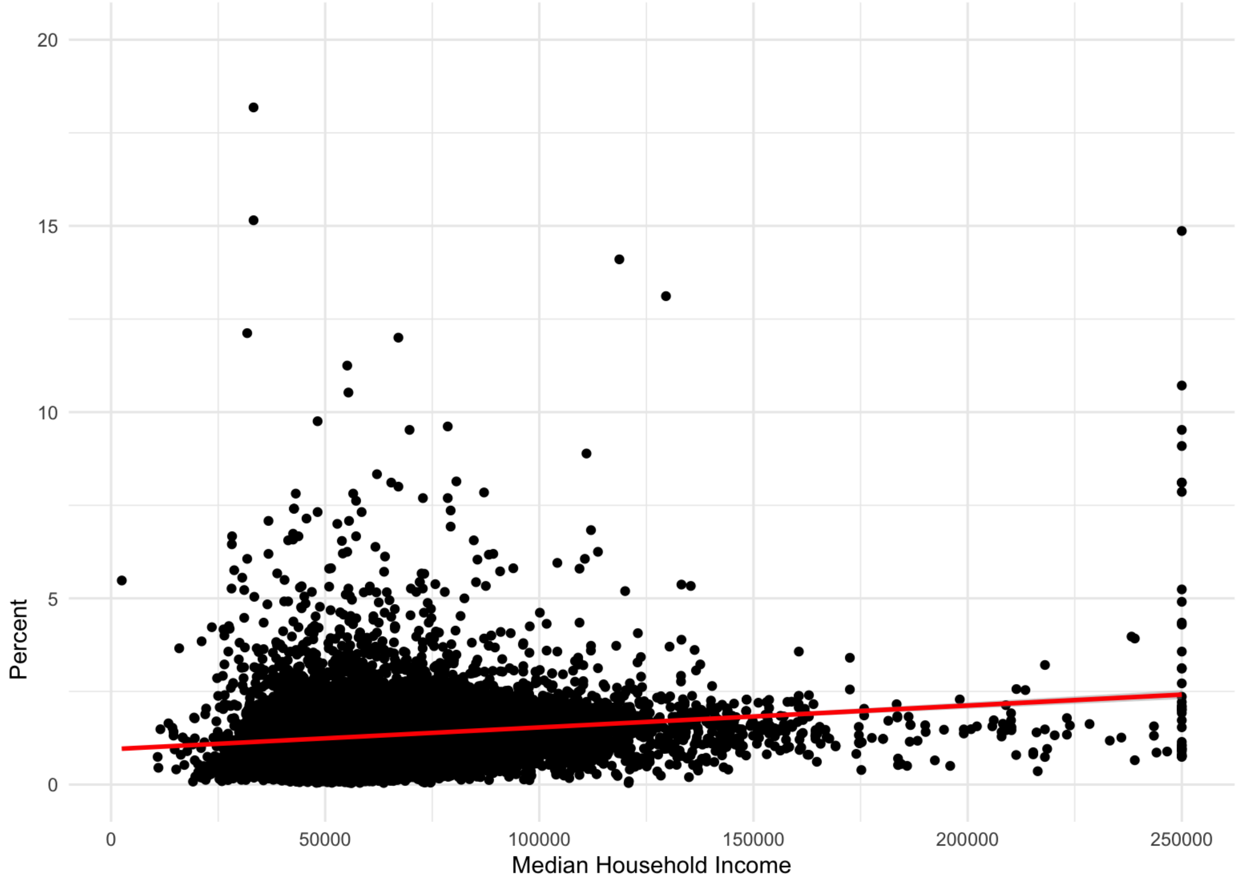


Scatterplot of the percent of adults with a medical cannabis certification for pain versus median household income in 2024 at the ZCTA level (r(1,606) = 0.0278, p = 0.264)

**Supplementary Fig. 12.**

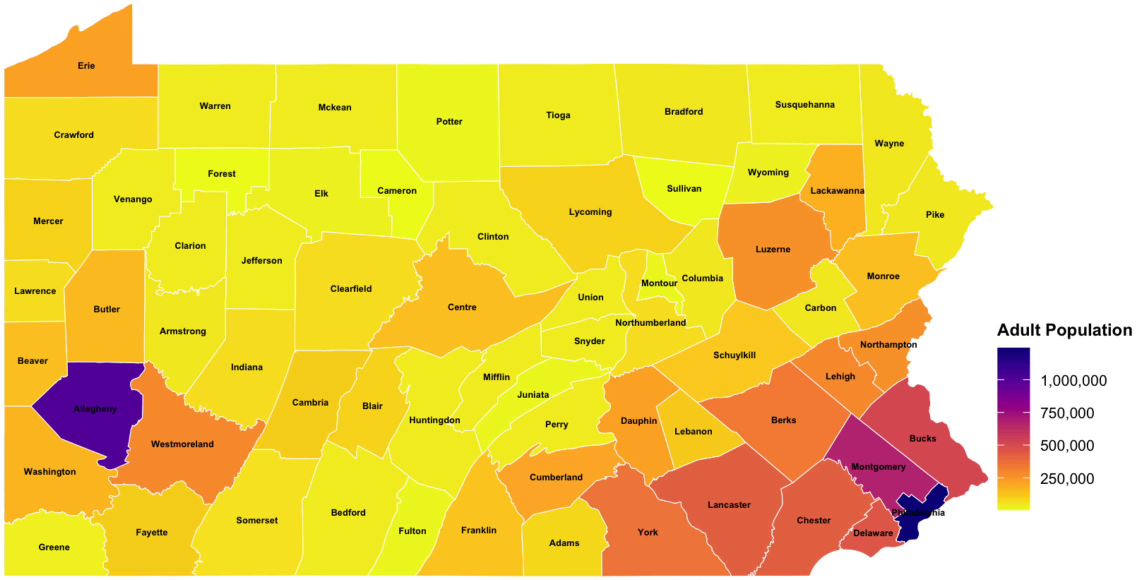


Heat map of the adult population in the counties in Pennsylvania. Over half (55.5%) of adults in Pennsylvania in 2024 resided in ten counties (Philadelphia, Allegheny, Montgomery, Bucks, Delaware, Lancaster, Chester, York, Berks, Lehigh).

**Supplementary Fig. 13.**

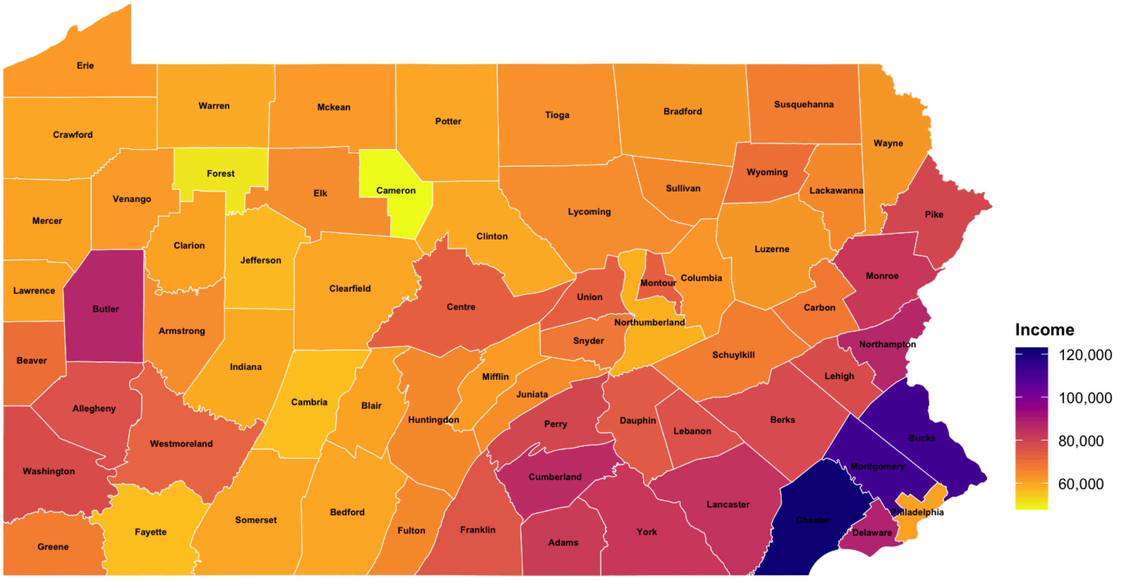


Heat map of the median household income in the counties in Pennsylvania in 2024. Pittsburgh is in Allegheny County while Philadelphia is in Philadelphia County.
